# Multi-ancestry Genome-Wide Association Meta-Analysis Identifies Novel Loci in Atopic Dermatitis

**DOI:** 10.1101/2024.06.17.24308897

**Authors:** Meritxell Oliva, Mrinal K. Sarkar, Michael E. March, Amir Hossein Saeidian, Frank D. Mentch, Chen-Lin Hsieh, Fanying Tang, Ranjitha Uppala, Matthew T. Patrick, Qinmengge Li, Rachael Bogle, J. Michelle Kahlenberg, Deborah Watson, Joseph T. Glessner, Lam C. Tsoi, Hakon Hakonarson, Johann E. Gudjonsson, Kathleen M. Smith, Bridget Riley-Gillis

## Abstract

Atopic dermatitis (AD) is a highly heritable and common inflammatory skin condition affecting children and adults worldwide. Multi-ancestry approaches to AD genetic association studies are poised to boost power to detect genetic signal and identify ancestry-specific loci contributing to AD risk. Here, we present a multi-ancestry GWAS meta-analysis of twelve AD cohorts from five ancestral populations totaling 56,146 cases and 602,280 controls. We report 101 genomic loci associated with AD, including 15 loci that have not been previously associated with AD or eczema. Fine-mapping, QTL colocalization, and cell-type enrichment analyses identified genes and cell types implicated in AD pathophysiology. Functional analyses in keratinocytes provide evidence for genes that could play a role in AD through epidermal barrier function. Our study provides new insights into the etiology of AD by harnessing multiple genetic and functional approaches to unveil the mechanisms by which AD-associated variants impact genes and cell types.

**Disclosure Statement:** BRG, MO, CH, KMS are employees of AbbVie. FT was an employee of AbbVie at the time of the study. JEG (University of Michigan) has received research support from AbbVie, Janssen, Almirall, Prometheus Biosciences/Merck, BMS/Celgene, Boehringer Ingelheim, Galderma, Eli Lilly, and advisor to Sanofi, Eli Lilly, Galderma, BMS, Boehringer Ingelheim. MKS, RU, MTP, QL, RW, JMK, LCT are employees of University of Michigan and have no funding to disclose. MEM, AHS, FDM, DW, JTG, HH are employees of the Children’s Hospital of Philadelphia and no funding to disclose. The design, study conduct, and financial support for this research were provided by AbbVie. AbbVie participated in the interpretation of data, review, and approval of the publication.

## MAIN

Atopic dermatitis (AD) is one of the most common chronic conditions, affecting 15-20% of children and 5-10% of adults worldwide^1,2^. AD is characterized as a pruritic rash primarily on the flexural areas of the arms and legs, and can vary widely in severity and presentation^3,4^. Both genetic and environmental factors can predispose individuals to impaired epidermal barrier function and inflammation that, along with itch, drive the vicious cycle of AD^5,6^. Genetic studies published to date support a role for genetic defects in innate and adaptive immunity resulting in Th2 skewing as well as defects in skin barrier function, most notably loss-of-function variants in filaggrin (FLG)^7,8^. AD heritability has been estimated from twin studies to be 75-80%^9–11^, and there are currently more than 90 published GWAS loci that explain about 5-15% of heritability^12–14^, suggesting additional AD genetic loci are to be discovered.

As GWAS data in diverse populations becomes available, the human genetics field has moved toward multi-ancestry approaches. Multi-ancestry GWAS meta-analysis across diverse populations can increase the power to detect complex trait loci when the underlying causal variants are shared between ancestry groups. Our study is motivated by previous reports and our own experience meta-analyzing European cohorts and the desire to incorporate diverse populations of Asian and African ancestry. Moreover, we know that AD affects populations worldwide, and diverse populations can expand our understanding of the genetic architecture of AD.

Here, we report a large-scale multi-ancestry GWAS of 56,146 AD cases and 602,280 controls as well as ancestry-specific analyses. The cohorts include individuals of European (EUR), Asian (ASN), African (AFR), and American (AMR) ancestries. In total, we report 101 genome-wide significant loci associated with AD, including 15 loci that have not been previously reported as associated with AD or eczema. We characterize novel genetic factors for AD across multiple ancestral populations and perform fine-mapping and colocalization to identify putatively causal genes at genome-wide significant loci. Cell-type enrichment analysis identifies the disease-relevant cell types implicated by the GWAS signal, including keratinocytes, in which we perform functional experiments. Our findings highlight newly associated loci and link genes in these loci to cell types in key AD mechanistic nodes.

## RESULTS

### Multi-ancestry and ancestry-specific GWAS meta-analyses identify novel AD loci

We performed GWAS meta-analysis on 12 cohorts comprising a total of 56,146 AD cases and 602,280 controls of European (EUR), Asian (ASN), African (AFR), American (AMR) and admixed ancestries (**Fig. 1**, **Supplementary Table 1**) to obtain ancestry-specific (EUR, EAS and AFR) and multi-ancestry (MULTI) GWAS summary statistics (Methods, **Supplementary Table 2**). Across all GWAS meta-analyses, we detected 101 genome-wide significant (P < 5e-08) non-overlapping loci (**Supplementary Table 3**), including 15 loci not reported as associated to AD or eczema to date (**Table 1**); we will refer to these as novel. Novel AD loci were identified in the two largest GWAS meta-analyses (MULTI and EUR) (**Fig. 2a**, **Table 1**). The 12 genome-wide significant loci identified in the ASN meta-analysis (**Extended Fig. 1a**) were located within the significant loci from the MULTI meta-analysis (**Fig. 2b),** however, comparison of the MULTI meta-analysis with and without the ASN cohorts confirms increased p-value significance for lead SNPs when the ASN samples are included (**Extended Fig. 1b**). No genome-wide significant loci at MAF > 0.01 were identified in the AFR GWAS meta-analysis despite the number of cases included (N = 7,063); corresponding GWAS signal was seemingly well-controlled (*λ*_gc_ = 1.02) but underpowered (**Extended Fig. 1, Supplementary Table 2**), which may be due, among possible factors, to genetic admixture and differential environmental exposure^15^.

**Figure 1.**
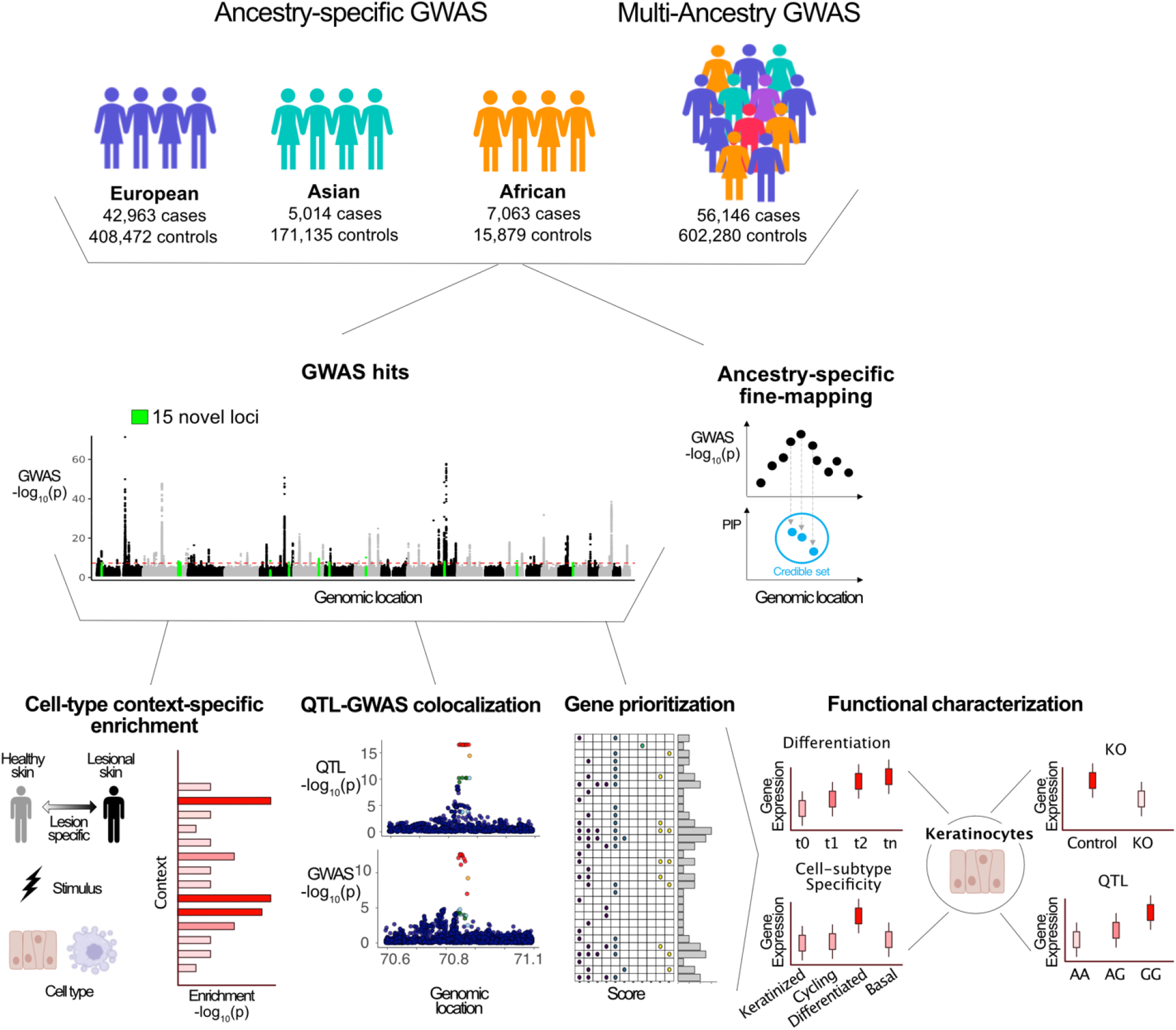
Scheme of data generation and analysis overview. Ancestry-stratified GWAS meta-analyses were utilized to identify and fine-map genome-wide significant GWAS loci for atopic dermatitis risk. GWAS signal was integrated with functional maps and evaluated for cell-type context-specific enrichment and QTL colocalization; genes were prioritized per GWAS locus. The expression of a subset of prioritized genes was functionally characterized in keratinocytes for differentiation, cell-subtype specificity, knock-out (KO) and Quantitative Trait Loci (QTL) signal. This figure contains illustrations from http://biorender.com.

**Figure 2.**
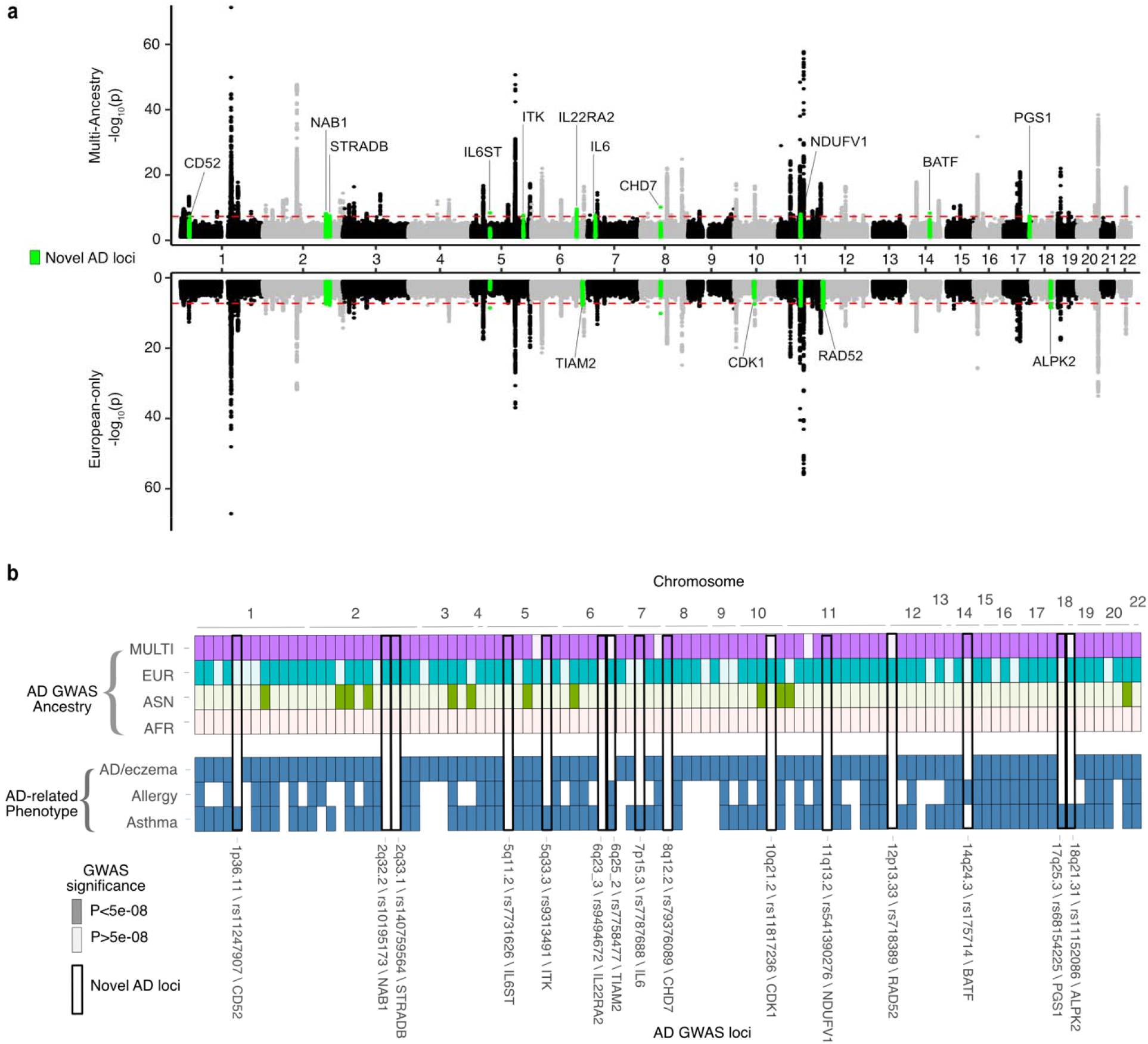
AD GWAS loci annotation by ancestry and AD-related phenotypes. **a**, P-values (y axis) derived from the multiple ancestry-combined (MULTI) (top panel) and the European-ancestry stratified (EUR) (bottom panel) GWAS meta-analysis are plotted by corresponding genomic coordinate (x axis). Novel associations are highlighted in green and annotated with the nearest gene. Associations that reached significance (P<5e-08) in both MULTI and EUR GWAS meta-analyses are annotated in the top panel; associations that reached significance (P<5e-08) in only the EUR GWAS meta-analysis are annotated in the bottom panel. **b**, Top panel illustrates the presence of significant (P<5e-08) AD GWAS loci (x axis) across ancestry-stratified and -combined GWAS meta-analyses (y axis); significant loci per ancestry endpoint are indicated with a darker color shade. Bottom panel illustrates overlap of significant AD GWAS loci with previously reported genome-wide significant (P<5e-08) GWAS loci for AD/eczema, allergy, and asthma phenotypes. Bold frame indicates the 15 novel AD GWAS loci reported herein, annotated with corresponding cytoband, lead variant and nearest gene.

**Table 1.**
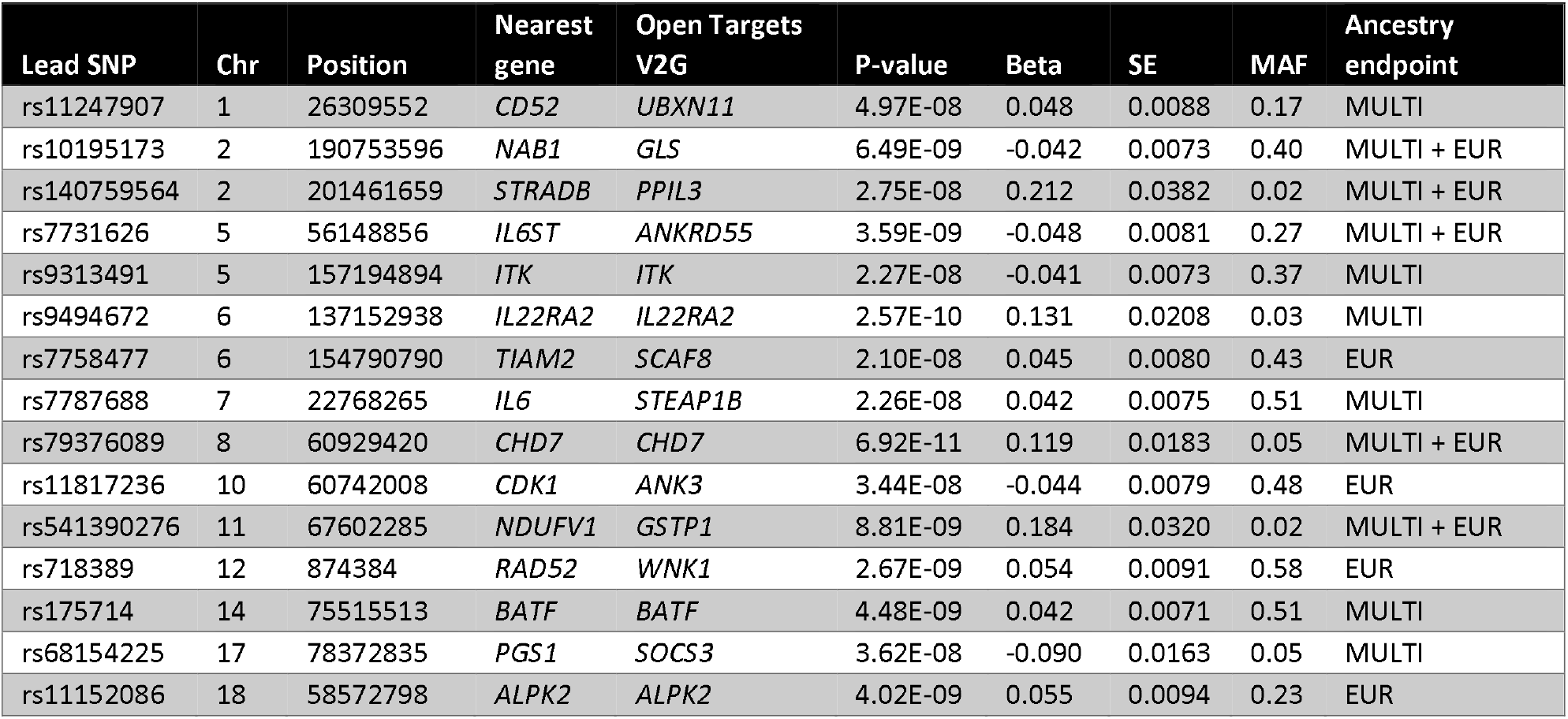
AD GWAS novel loci. Summary statistics and genomic context of the 15 AD GWAS loci not previously reported in eczema and AD GWAS studies (Methods).

We quantified heritability - the proportion of phenotypic variance explained by genetics - in the European cohort to be 9.67% on a liability scale (Methods). This estimate agrees with previous reports ^11,13^, but is substantially smaller than the 75-80% estimated heritability derived from twin studies^9–11^. This may be due to several factors, including contribution from genetic loci not captured by the European ancestry meta-analysis, rare variants with large effect sizes not captured by genotyping arrays, complex gene-gene or gene-environment interactions important to the genetic architecture of AD, or­ overestimation of twin heritability^16^.

To better understand the relationship between the AD-associated loci identified herein and previously published results on AD-related phenotypes, we assessed overlap with reported genomic associations for atopic march phenotypes other than AD: eczema, allergy, and asthma (**Methods**). Out of the 101 loci identified in our analysis, we observe that the majority (93/101) overlap reported atopic-march associated loci (**Fig. 2b**), including 7 out of the 15 novel loci not previously reported to be associated with AD. The overlap supports the known shared genetic architecture of AD with other atopic-march phenotypes^17,18^ and pinpoints additional contributing loci. Of the 8 novel loci not previously reported in AD/eczema, allergy, or asthma GWAS, only 1 locus - rs7731626 (5q11.2) - has reported disease associations in GWAS catalog. This locus is associated with a range of autoimmune phenotypes such as rheumatoid arthritis^19^, inflammatory disease^19^, multiple sclerosis^20^, and type 1 diabetes^21^. Combined, these results suggest that we have replicated genetic loci that play a role in atopic disease and identified novel loci that expand the current knowledge of the genetic architecture of AD.

### Cell-type enrichment confirms key AD mechanistic nodes

To identify the disease-relevant cell types impacted by the identified AD relevant loci, we integrated GWAS signal with epigenetic and single-cell transcriptomic annotations. LD score regression (LDSC-SEG) was used to identify genomic annotations enriched for genetic trait heritability in the EUR GWAS, and bulk ATAC-seq data for cells isolated from peripheral blood from healthy donors (GSE118189, **Methods**)^22^. AD GWAS variants are enriched primarily in open chromatin of T-cell populations such Th1, Th2, Th17, and Treg cells. These findings support the known pathobiology of T-cell driven inflammation in AD^23,24^ (**Fig. 3a**).

**Figure 3.**
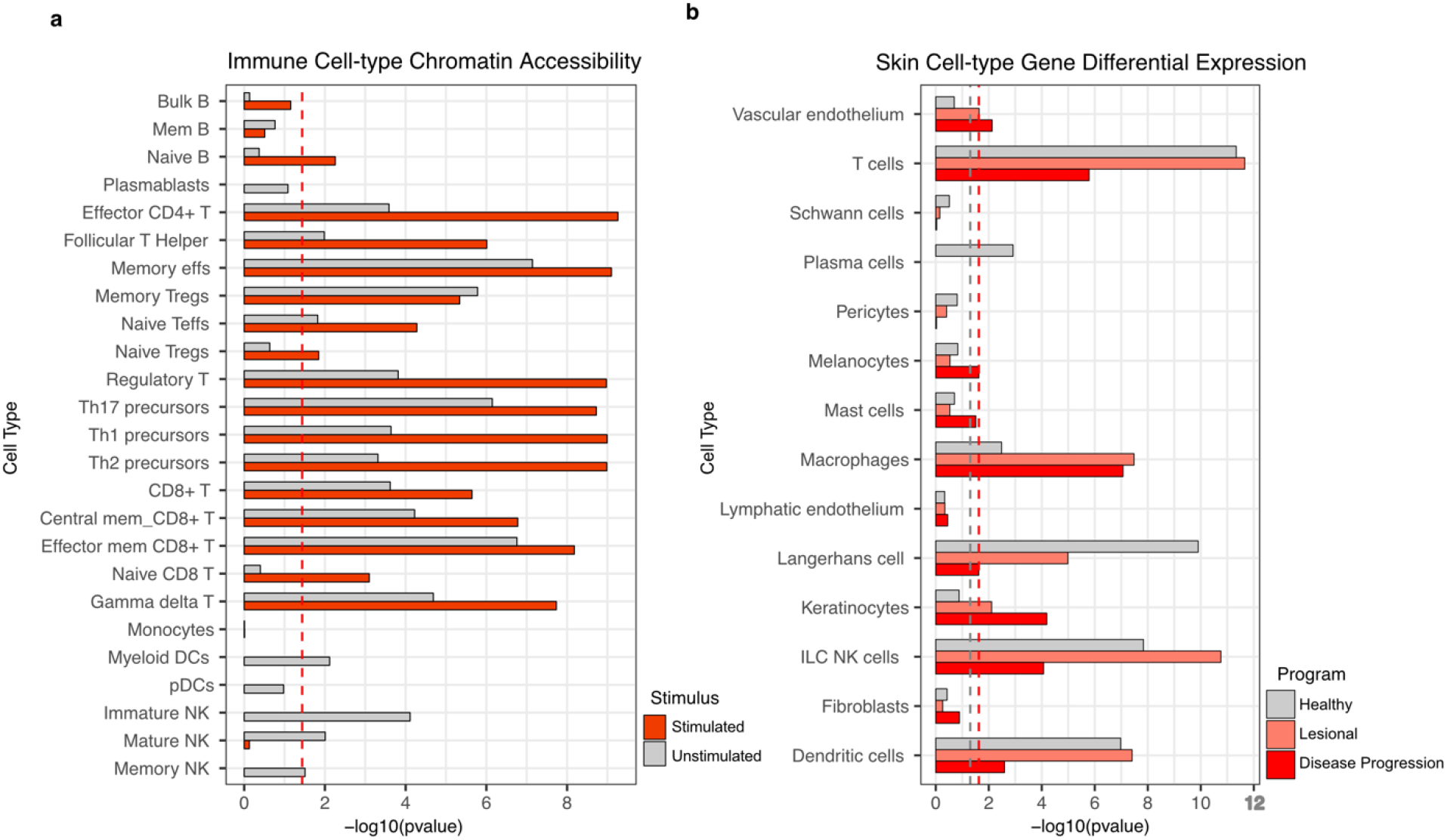
Cell-type and context-specific enrichment of AD GWAS signal. **a**, AD GWAS heritability enrichment (x axis) in accessible chromatin per immune cell type (y axis), derived from LDSC-SEG analyses of stimulated (red) and non-stimulated (grey) isolated immune cell ATAC-seq data annotations (Methods). Red dashed line corresponds to FDR < 0.05 threshold. **b**, AD GWAS signal enrichment (x axis) in differentially expressed genes per skin cell type (y axis), derived from MAGMA analyses of healthy and AD-affected skin sc-RNAseq gene expression (Methods). Differential expression corresponds to healthy (grey), lesional (pink) and AD progression (red) programs. ‘Healthy’ and ‘Lesional’ programs are defined by genes highly expressed in a particular skin cell type compared to others. Disease progression program is defined as differential expressed genes between cells of the same type in AD-lesional relative to healthy tissue. Red and grey dashed line corresponds to FDR < 0.05 and P < 0.05 thresholds, respectively.

To explore disease-relevant cell types in skin, we utilized MAGMA to evaluate the enrichment of GWAS signal in skin cell types from the Human Cell Atlas skin dataset^25^, a single cell RNAseq dataset (sc-RNAseq) derived from healthy skin tissue (H) and AD lesional skin tissue (LS) (**Methods**). Cell-type-specific gene programs (differential gene expression in one cell type compared to other cell types in H or LS tissue) and disease progression (DP) programs (differential expression between cells of the same type in LS *vs*. H tissue) were constructed for 14 cell type categories to test for enrichment of GWAS signal (**Methods**). For cell type and DP programs, strong enrichment was identified in lymphocytes, including T-cells, NK cells, and innate lymphoid cells (ILCs), as well as macrophages and dendritic cells. These findings underscore the contributions of both innate and adaptive immune cell types in the inflammatory node of AD biology. Enrichment in keratinocytes pinpoints another key mechanistic node in AD biology, the skin barrier. This signal is significantly enriched in LS skin from AD patients and in the DP program, but not in healthy skin (**Fig. 3b**). Refinement of the keratinocyte cluster into defined sub-populations shows strongest enrichment in differentiated keratinocytes in LS and DP programs followed by undifferentiated and proliferating keratinocytes in DP program and inflammatory differentiated KCs in LS program (**Extended Fig. 2**). The specific keratinocyte enrichment patterns underscore the role for AD GWAS implicated genes in epidermal differentiation and barrier function in the upper layers of the epidermis.

To investigate the genetic contribution of the AD GWAS signal to the cell types identified in **Fig. 3b**, we clustered the gene program scores for the 146 genes with MAGMA p-value < 2.7e-06 for the 14 cell-type clusters and identified distinct clustering by immune and non-immune cell types in the skin (**Extended Fig. 3**). Additionally, the DP cell types frequently clustered separately from the H and LS cell types, indicating different roles for AD GWAS genes in H and LS states compared to DP. An exception is for the keratinocytes and melanocytes, where the LS and DP programs cluster together. Enrichment of the GWAS signal in keratinocytes and clustering of LS and DP keratinocyte gene programs supports further exploration of AD GWAS genes influencing keratinocytes and the role in barrier function.

Integration of functional annotations identifies putative causal genes at AD GWAS loci

To identify putative causal genes and variants at each locus, we performed the following analyses. First, we performed fine-mapping on 78/101 loci that reached genome-wide significance (P<5e-08) in EUR (75/78) or ASN (11/78) meta-GWAS analyses (excluding MHC region). We identified 13 credible set variants annotated as moderate or high impact in 9 genes (**Supplementary Table 4**) including 3 coding variants in the *FLG*, *TESPA1*, and *NLRP10* loci with posterior inclusion probability (PIP) >0.9 (**Supplementary Table 4**). Notably, for the fine-mapped variants in *TESPA1* (rs183884396, PIP = 0.99) and *NLRP10* (rs59039403, PIP = 1), the minor alleles are enriched in specific ancestral populations. *TESPA1*_rs183884396-G allele frequency is >14x enriched in the Finnish population compared to non-Finnish Europeans and *NLRP10*_rs59039403-G has an allele frequency in East Asian populations of 12.4% compared to <0.1% in EUR populations, demonstrating the value of cohorts from diverse ancestries to identify genetic signal for AD. Second, we identified coding variants in LD with the lead variants from the 3 GWAS meta-analyses performed with genome-wide significant results (P < 5e-8), excluding the HLA region (r^2^ > 0.6, MULTI, EUR, EAS samples in 1KG Phase 3; **Methods**) and annotated the coding variants predicted to impact gene function. Twenty-seven coding variants in 19 genes were identified across the MULTI, EUR and ASN meta-analyses (**Supplementary Table 5**). The identified genes include reported causal or putatively causal AD genes such as *FLG*^26^ and *NLRP10*^26^ as well as genes with reported eczema, dermatitis or immunodeficiency phenotypes in OMIM, *SIK3*, *IL7R* and *RTEL1*, respectively. Combined, the genes with coding variation in the fine-mapped credible sets and in LD with the GWAS lead variants identify potentially causal AD genes.

Next, we performed colocalization with Quantitative Trait Loci (QTLs) to infer molecular consequences of the AD GWAS variants. We compiled and harmonized a QTL catalog of 297 full summary statistics of expression (eQTL), splicing (sQTL), protein (pQTL) and DNAm (mQTL) maps maximizing the inclusion of immune cell types relevant to AD (**Supplementary Table 6**). In total, we identified 3,195 colocalizations (PP4>0.75) across the majority (86/101) of AD-associated loci including 12/15 novel loci (**Supplementary Table 7, Fig. 4a**). While for most (85%, 73/86) QTL-associated loci, colocalization(s) at gene expression level (eQTL) were identified, 13/86 loci lacked eQTL associations but were supported by other molecular phenotypes (**Extended Figure 4)**. Among those, we identified genes related to pathways relevant to AD, e.g. IL-22 genes, and less characterized genes, e.g. *CLEC16A*, uncharacterized in AD but reported to be a master regulator of autoimmunity^27^. It has been shown that multiple regulatory effects for the same gene often mediate the same complex trait associations, and that QTLs derived from different molecular phenotypes have an independent contribution to complex traits ^28,29^. We quantified QTL support per gene (**Fig. 4b**) and identified *CRAT, IL6R, IL7R* and *INPP5D* as supported by all QTL molecular phenotypes, and by multiple cells/tissue QTL endpoints. While CRAT locus GWAS lead variant is located in an intronic region of the uncharacterized protein-coding gene ENSG00000235007, the AD risk allele rs1107329-C is associated (P=6.9e-44, β=0.25) with increased CRAT protein abundance in plasma and increased levels of 2-methylmalonylcarnitine^30^ (**Fig. 4c**). CRAT encodes carnitine O-acetyltransferase; CRAT transcript levels and acetylcarnitines are reported to be altered in skin^31^ and serum of AD patients^32^, respectively. These results indicate that rs1107329-C increased AD risk may derive from genetic impact on carnitine metabolism.

**Figure 4.**
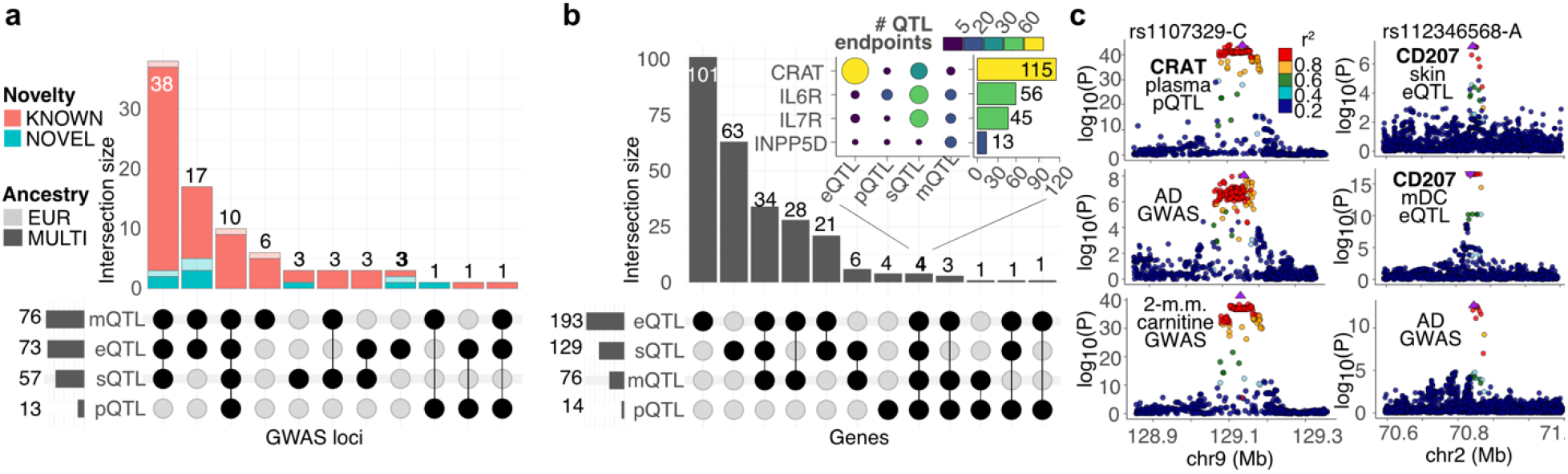
AD GWAS-QTL colocalization results. **a**, Number of AD GWAS loci with identified QTL colocalizations, stratified by QTL molecular phenotype and GWAS ancestry and novelty endpoints. **b**, Number of genes with identified QTL colocalizations, stratified by QTL molecular phenotype type. Inlet heatmap represents the number of QTL endpoints per QTL type (x axis) for each of the four genes supported by all QTL types (y axis); barplot shows aggregated cross-QTL support per gene. **c**, Genotype-phenotype association p-values of the CRAT and CD207 locus. Left panels illustrate CRAT pQTL signal in ARIC EA plasma (top) and GWAS signal for AD (middle) and GWAS signal for increased levels of 2-methylmalonylcarnitine (bottom). Right panels illustrate CD207 eQTL signal in GTEX skin not exposed to sun (top), ImmuNexUT myeloid dendritic cells (mDC) (middle) and GWAS signal for AD (bottom). Lead GWAS variant mapped to AD risk allele is typed and illustrated by a triangle-shaped point pointing upwards, indicating allele association with positive phenotype effect. Linkage disequilibrium between loci is quantified by squared Pearson coefficient of correlation (r2). P-values correspond to nominal GWAS and QTL associations, derived from multiple regression two-sided t-tests.

The usage of diverse cell and tissue QTL endpoints can aid the prioritization of cell of origin for a candidate gene. For example, we identified *CD207* colocalization instances exclusively in skin and myeloid dendritic cell (DC) eQTL endpoints, with the AD risk allele rs4852714-A associated with decreased and increased expression of *CD207* in skin and DCs, respectively (**Fig. 4c**). In skin, *CD207* is exclusively expressed in Langerhans cells (LCs), which are epidermal resident DCs of the myeloid lineage. These results pinpoint skin-resident dendritic cells as the causal cell of origin type of rs4852714-A increased AD risk and demonstrate the need of skin QTL maps at cell-type resolution to confidently assess the effect directionality of AD risk alleles on impacted genes in causal cell-type contexts.

Finally, to prioritize genes at each locus, we scored each candidate gene within the locus by aggregated support from multiple lines of evidence, including variant-to-gene predictions, QTL evidence, coding variant genes, AD phenotype annotations (**Methods**, **Supplementary table 8, Extended Fig. 5**). Top-scoring genes in novel loci include *ITK* and *BATF* (**Fig. 5a**). *ITK* encodes IL-2 inducible T-cell kinase, it colocalizes exclusively in T-cells (**Fig. 5c**) and is upregulated in AD lesional skin. Autosomal recessive mutations in *ITK* cause Lymphoproliferative Syndrome 1, a primary immunodeficiency characterized by early childhood Epstein-Barr virus associated immune dysregulation manifesting in lymphoma and autoimmune disorders. *BATF* encodes basic leucine zipper ATF-like transcription factor, it colocalizes in CD8+ memory T-cells and is upregulated in AD lesional skin. AD GWAS associations with the BATF gene family member *BATF3* have been reported^33^. In mice, the Batf/Batf3 interaction controls Th2-type immune response through regulation of IL-4 production^34^. Given their role in immune response, T-cell function, and reported links to immune diseases, *ITK* and *BATF* may play a role in AD biology and warrant further investigation.

**Figure 5.**
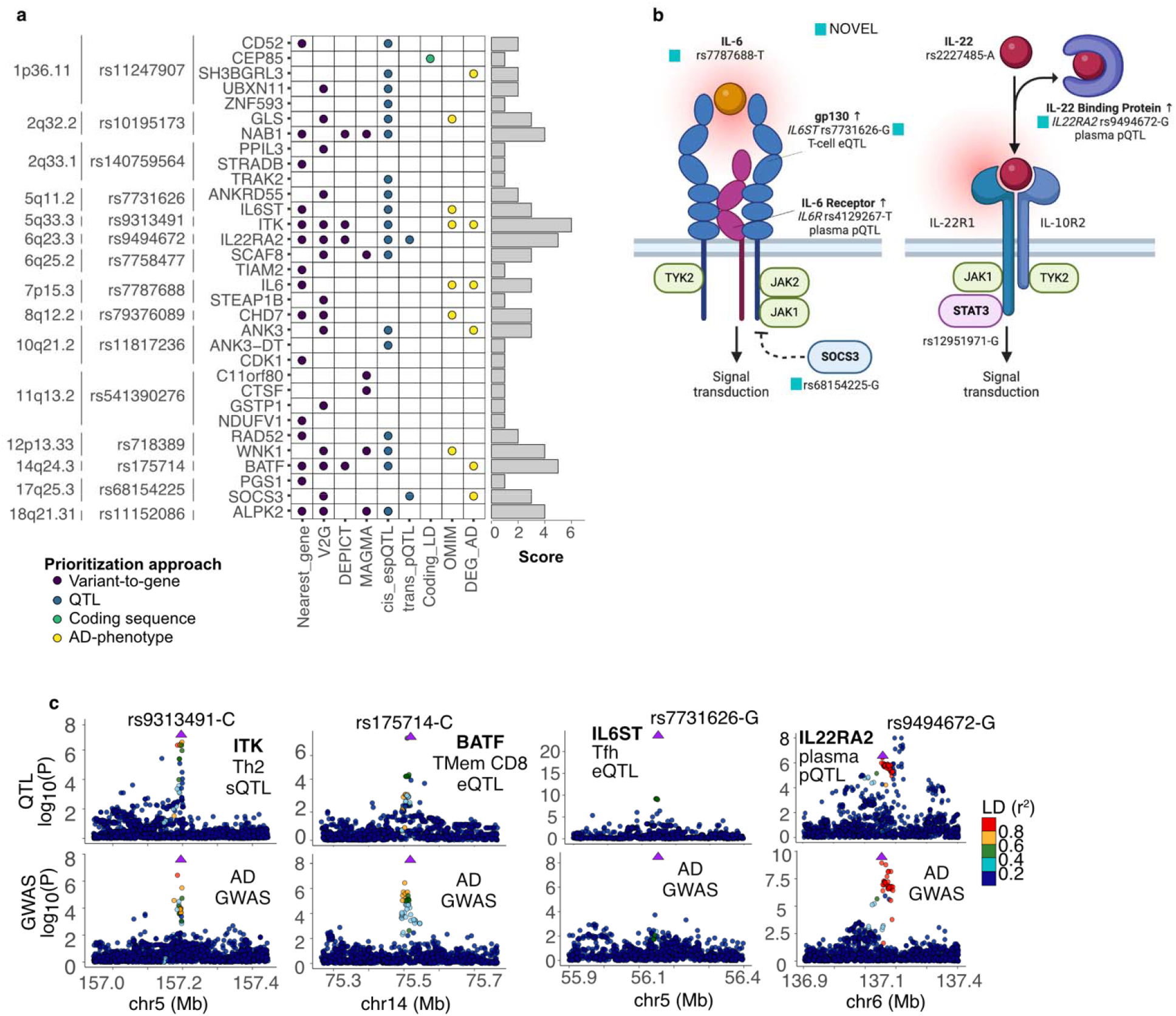
Prioritized candidate genes at novel loci. **A,** Supporting evidence by candidate per GWAS locus. **B**, Scheme of ligand-receptor and proximal components of IL-6 and IL-22 signaling pathway. Corresponding expression and protein QTL associations mapped to the lead AD GWAS variant risk allele are shown. The figure was created using BioRender. **C**, Genotype-phenotype association p-values of the IL6ST, IL22RA2, ITK and BATF locus. First panel illustrates ITK ENST00000522616 sQTL signal in T helper 2 cells (Th2), second panel illustrates BATF eQTL signal in CD8+ memory T-cells, third panel illustrates IL6ST eQTL signal in T follicular helper cells (Tfh), forth panel illustrates IL22RA2 pQTL signal in plasma. For all panels, bottom part illustrates GWAS signal for AD. Lead GWAS variant mapped to AD risk allele is typed and indicated by a triangle-shaped point, pointing upwards or downwards for positive or negative effect size, respectively. Linkage disequilibrium between loci is quantified by squared Pearson coefficient of correlation (r^2^). P-values correspond to nominal GWAS and QTL associations, derived from multiple regression two-sided t-tests.

Among high-scoring, prioritized genes we found multiple instances of receptor–ligand interactions for circulating cytokines and TNF-superfamily members, including genes involved in IL-6 and IL-22 signaling pathways (**Fig. 5b-c**). The variant rs7731626 is located within an intron of *ANKRD55* and colocalizes with T-cell eQTLs for both *ANKRD55* and *IL6ST,* which encodes IL-6 receptor complex protein gp130. We, and others^35^, prioritize *IL6ST* as the putative causal gene, linking the locus to the IL-6 signaling pathway that plays a key role in autoimmune and chronic inflammatory diseases. We identified additional IL-6 pathway genes – *IL6*, *IL6R* (IL-6 receptor), and *SOCS3* (JAK/STAT inhibitor) – as prioritized candidates for corresponding AD GWAS loci. In the IL-22 pathway, we identified both *IL22* and *IL22RA2* as prioritized AD candidates. *IL22RA2* is the gene prioritized at the most significant novel AD GWAS locus (P=2.57e-10) and has support from both *cis* and *trans* pQTL signals in plasma. The *trans IL22RA2* pQTL variant rs4265380 is located at the prioritized gene *RUNX3* locus. *RUNX3* has been previously associated with psoriasis^36^ and may modulate frequency of Th17 and Th22 cells^37^; *RUNX3* and *IL22RA2* interactions have been reported to be involved in macrophages IL-22 mediated intestinal inflammatory response in mouse leading to colitis^38^. The prioritization approach captures known and uncharacterized AD molecular associations, aids the prioritization of putative AD-causal genes and cellular contexts, and provides insight into gene sets and pathways contributing to AD pathobiology.

### Integration of colocalization and functional assays identifies keratinocyte-specific AD-linked genes

Skin barrier defects, a key feature of AD, primarily affect keratinocytes^39^, and we observed AD genetic signal enrichment for genes differentially expressed in keratinocytes from AD patients (**Fig. 3**). Hence, we hypothesized that a fraction of the colocalized genes impact AD by altering keratinocyte-specific gene expression programs and mechanisms. To explore our hypothesis, we selected 22 colocalized genes associated with keratinocyte-specific signatures (**Methods, Supplementary Table 9**). We defined this set as “AD keratinocyte-linked gene candidates” and profiled them by four complementary functional assays. Firstly, to evaluate their enhanced expression in keratinocyte populations, we generated sc-RNAseq profiles of epidermal cells from seven human body sites (**Methods**, **Extended Fig. 6**). Secondly, to investigate their involvement in keratinocyte differentiation, we generated a three-dimensional epidermal model and generated bulk-RNA-Seq profiles from seven differentiation timepoints (**Methods, Extended Fig. 7**). Thirdly, we explored their response to the AD-relevant cytokine pathways IL-13 and IL-22^40^, considered the two major effector cytokines in AD pathogenesis^41,42^, by silencing each gene in keratinocyte models and characterizing the expression of cytokine IL-22 and IL-13 pathway proxy genes under different treatment conditions: no treatment, stimulation with IL-22, IL-13, or both (**Methods**). Finally, to assess whether AD risk alleles affect transcript abundance in keratinocytes in inflammation contexts, we generated a total of 8 *cis* eQTL maps from keratinocyte cell lines derived from 50 human subjects; cells were untreated or treated with cytokine treatments IFNa, IFNb, IL-13, IL-17, IL-4, IL-17+TNFa and TNFa (**Methods**).

We observed that, compared to non-prioritized candidates (**Extended Fig. 8)**, per-locus prioritized AD keratinocyte-linked genes (**Fig. 6)** tend to yield significant effects across assays. Considering differential expression by keratinocyte cell population, we identified *CEBPA* as enhanced in differentiated keratinocytes, *AQP3* and *RGS14* in non-keratinized keratinocytes, and *RORA* and *ANK3* in keratinized populations. In epidermal models, *CEBPA, AQP3*, *RORA* and *ANK3* were identified as strongly positively associated with keratinocyte differentiation (**Fig. 6**). While cytokine pathway signal differs by treatment, all tested prioritized candidates – except *RGS14* – show nominally significant (t-test, P<0.01) signal in at least one treatment condition (**Extended Fig. 9b**). Despite the limited power of the keratinocyte eQTL maps, all tested prioritized candidates show nominally significant (t-test, P<0.05) eQTL signal in at least one stimulated keratinocyte source, confirming that AD risk alleles impact candidate genes expression in keratinocytes (**Fig. 6**, **Supplementary Table 9).**

**Figure 6.**
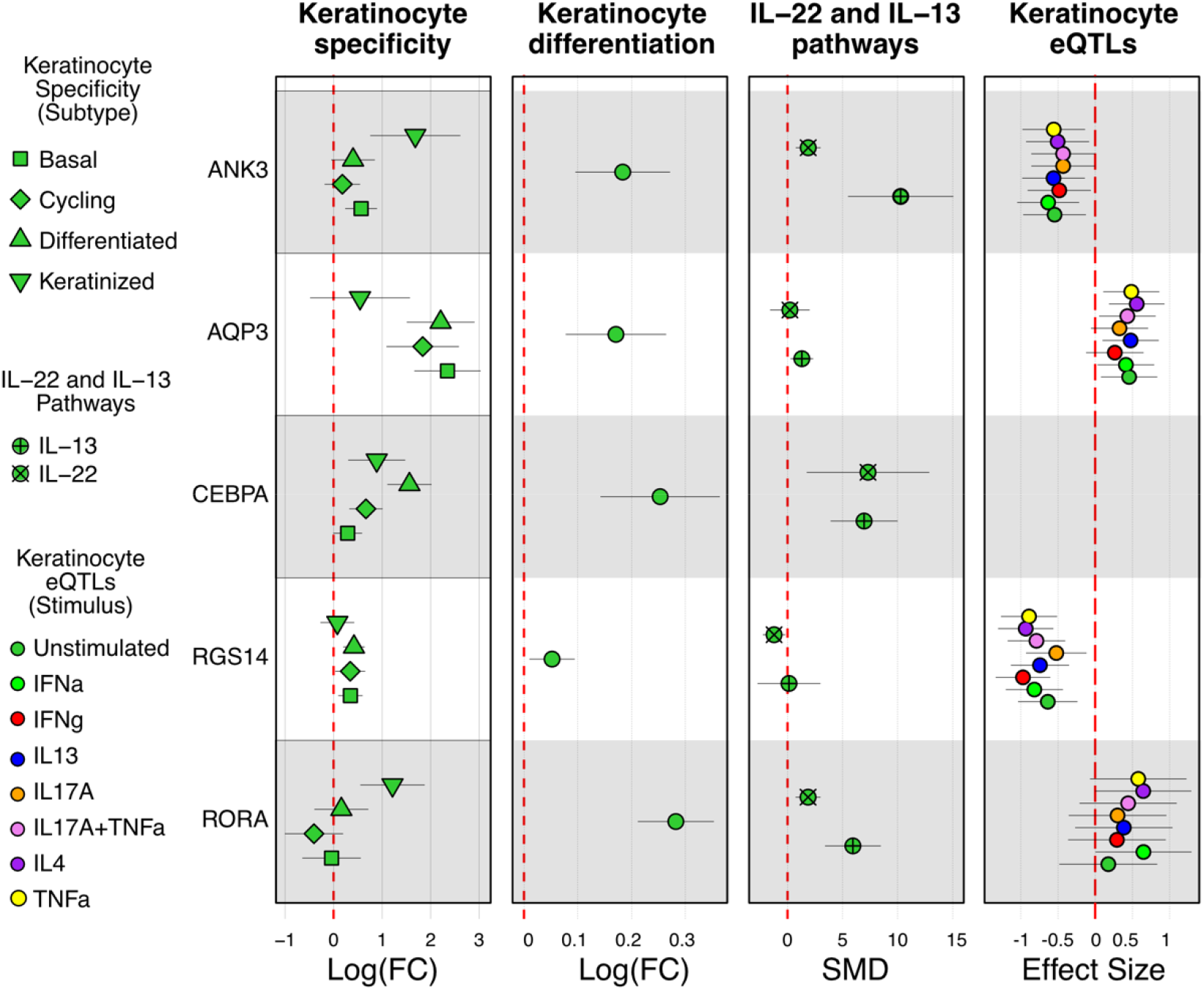
Functional characterization of prioritized AD keratinocyte-linked gene candidates. **1st panel**, Keratinocyte subtype differential expression (x axis) by gene (y axis). Differential expression values correspond to expression log fold change (FC) between keratinocyte and non-keratinocyte cells, mean-averaged across body sites, and whiskers represent the standard deviation of the mean. **2nd panel**, Differential expression (x axis) of gene (y axis) as a function of epidermal raft differentiation. Differential expression values correspond to log expression fold change as a function of differentiation timepoints (Methods), and whiskers represent the 95% confidence interval of the value. **3rd panel**, Differential expression of pathway (x axis) proxy genes by candidate gene (y axis). Differential expression values correspond to the standardized mean difference (SMD) of the expression of genes that are proxies of IL-22 and IL-13 pathways between the presence or absence of siRNA targeting corresponding gene candidate. Whiskers represent the 95% confidence interval of the value. SMD values were derived from across-proxy-genes per-pathway meta-analysis (Methods). **4th panel**, Keratinocyte eQTL effect size (x axis) by candidate gene (y axis), stratified by keratinocyte stimulus. Effects are mapped to AD risk alleles of the corresponding GWAS locus lead variants. Whiskers represent the 95% confidence interval of the value. CEBPA effect size is not shown due to insufficient AD risk allele frequency (Methods).

Combined, resources generated herein provide valuable mechanistic insights for uncharacterized AD genomic associations. One example is the novel *ANK3* locus. *ANK3* is known to play a role in neurodevelopmental disorders^43^, but its role in atopic dermatitis has not yet been described. We hypothesize that *ANK3* could be associated to AD immunopathogenesis by playing a role in keratinocyte-mediated inflammatory programs, as we observe that *ANK3* expression is enhanced in keratinized populations (granular layer), and the corresponding transcript silencing increases IL-13 pathway proximal gene expression in IL-13+IL-22 co-stimulated keratinocytes **(Fig. 6, Extended Fig. 9**). The AD risk allele rs11817236-A may impact *ANK3* in a cell-type specific manner; we observed decreased expression in keratinocytes (**Fig. 6**), but increased expression is reported in specific T-cell populations^44^. Together, these results highlight the utility of performing a comprehensive QTL-GWAS colocalization approach and integrating this data with cell-type relevant functional assays, to elucidate the potential mechanism by which genes not previously linked to AD could play a role in the disease by altering keratinocyte function.

## DISCUSSION

We present a large-scale, AD GWAS meta-analysis leveraging multi-ancestry cohorts, including N=13,183 non-European subjects affected by AD. We identified 101 loci associated with AD, including 15 novel loci not previously reported. While the inclusion of subjects from diverse ancestry backgrounds contributed to the overall multi-ancestry GWAS meta-analysis signal, we did not identify novel ancestry-specific GWAS loci in non-European cohorts. Fine-mapping of the ancestry-specific EUR and ASN GWAS meta-analyses identified variants with high posterior probability of being causal, including missense variants in *TESPA1* and *NLRP10* enriched in Finnish and East Asian populations, respectively. These findings support that larger AD cohorts in underrepresented non-European populations are required to increase the power to detect genome-wide significant variants, and that fine-mapping of ancestry-specific AD GWAS signal can identify putatively causal genes and variants. Furthermore, previous studies of AD and asthma in African Americans suggest that applying a stricter phenotype definition may aid discovery of novel signal^45,46^, and existing differential AD-triggering environmental exposures^15^ may contribute to explain the higher prevalence of AD for this community that is seemingly not explained by genetics^47^. To improve the characterization of AD genetic architecture, future efforts should not only continue the diversification and expansion of AD GWAS cohorts, but also focus on the refinement of phenotype definition and control of environmental exposure.

Exploring the genetic relationship between AD and atopic march phenotypes may help us to better understand the genetic architecture of disease. Atopic dermatitis often precedes the development of atopic march, defined as disease progression of AD to asthma and allergic rhinitis, which is associated with more severe and persistent disease^17^. We found that most of the identified AD loci overlap with previously reported atopic march GWAS loci, supporting the largely shared genetic architecture across atopic diseases. Both the replicated and novel AD loci highlight genomic regions that expand the understanding of AD biology.

Integration of multi-omic QTL sources from diverse biotypes to prioritize disease causal genes comprises the most extensive QTL-GWAS colocalization effort for AD genetic risk signals to date. Inclusion of complementary molecular phenotypes with cell-type specific QTL sources enabled the identification of AD-gene links otherwise missed. To prioritize genes at the novel and known loci, we leveraged multiple, complementary variant-to-gene approaches and functional annotations to derive a prioritization score. We focused on the biological interpretation of candidate genes at novel loci (for example *ITK* and *BATF*) and featured multiple prioritized genes in the IL-6 and IL-22 pathways, including *IL6ST*, *IL22RA2* and *SOCS3*, located in novel AD loci.

Cell-type enrichment analysis identified immune cells, particularly T-cells, as the top enriched cell type in the AD genetic signal, supporting the known pathobiology of T-cell driven inflammation in AD. Additionally, enrichment analysis in skin sc-RNAseq identified keratinocytes as a significant cell-type contributor. To our knowledge, this is the first report of AD genetic signal enrichment specific to keratinocytes, and supports the known role for skin barrier alterations in AD. To functionally characterize the keratinocyte signal on a selected gene candidate set, we generated a comprehensive array of skin and keratinocyte functional assays, including a cross-body epidermal sc-RNAseq atlas, keratinocyte multi-timepoint differentiation expression data, keratinocyte-derived eQTL maps and KOs across AD-relevant stimulation contexts in interleukin pathways. The integrated cross-assay results pinpoint *RORA, CEBPA, AQP3,* and *ANK3* as strong AD-linked candidates and highlight keratinocyte-subtype and context specificities. Importantly, the array of unique keratinocyte-derived resources generated herein can be further utilized by the scientific community to better understand AD pathobiology linked to the disruption of the epidermal barrier.

In conclusion, we leveraged multiple genetic and functional approaches to understand the mechanisms by which AD associated variants impact genes and cell types. We identified novel AD susceptibility loci, prioritized potentially causal genes, and pinpointed cellular contexts that contribute to the genetic architecture of AD. The provided resources can be utilized to further characterize the contribution of genetic signal to AD pathobiology and enable future efforts to identify AD-associated genes that may be transferrable into clinically actionable targets for atopic dermatitis.

## METHODS

### Study Populations

In this study, we included genetic and phenotype data from FinnGen, UK BioBank, and Children’s Hospital of Philadelphia and previously published GWAS from EAGLE consortium and BioBank Japan. The detailed information for each study is described below.

#### FinnGen

The FinnGen research project (www.finngen.fi) was launched in 2017 with an aim to improve human health through genetic research. The project combines genome information with digital health care data from national registries: the genotype data are linked to national hospital discharge, death, cancer, and medication reimbursement registries using the national personal identification numbers. Participants in FinnGen provided informed consent for biobank research on basis of the Finnish Biobank Act. Alternatively, separate research cohorts, collected before the Finnish Biobank Act came into effect (in September 2013) and the start of FinnGen (August 2017) were collected on the basis of study-specific consent and later transferred to the Finnish biobanks after approval by Fimea, the National Supervisory Authority for Welfare and Health. Recruitment protocols followed the biobank protocols approved by Fimea. The Coordinating Ethics Committee of the Hospital District of Helsinki and Uusimaa (HUS) approved the FinnGen study protocol (number HUS/990/2017). In the current analysis, we included 166,390 Finnish participants from FinnGen Data Freeze 10. Cases were obtained from ICD-8/9/10 diagnosis codes for atopic dermatitis, excluding other forms of dermatitis, from inpatient, outpatient or primary care registries and we required that a case also had a prescription code for AD medications from purchase or reimbursement registries. Controls excluded individuals with any dermatitis ICD codes or AD comorbidities (asthma and allergic rhinitis). The association analysis included 20,115 cases and 146,275 controls. GWAS was performed in the FinnGen Sandbox using the Scalable and Accurate Implementation of GEneralized mixed model (SAIGE v 0.36.3.2) including sex, age, genotyping batch and the first 10 genetic principal components. SNPs with minor allele count (MAC) > 5 and imputation quality score > 0.6 were kept in the association analysis.

#### UK Biobank

The UKBB is a large and population-based prospective cohort of approximately 500,000 participants aged 40-69 years recruited between 2006 and 2010 in the United Kingdom. This research was carried out using the UK Biobank resource under application number 26041. For the European ancestry analysis, we only included participants with European ancestry defined as Caucasian by the UKBB Field 22006. Atopic dermatitis cases were defined using ICD-9/10 codes for atopic dermatitis, excluding other forms of dermatitis, from hospital in-patient data and primary care and also had a prescription code for AD. Controls excluded individuals with any dermatitis ICD-9/10 codes or AD comorbidities (asthma and allergic rhinitis). The UKBB European ancestry association analysis included 10,470 cases and 210,720 controls.

For individuals of African and Central South Asian, we utilized the Pan-UK Biobank project (pan.ukbb.broadinstitute.org) assignment of UKBB participants to 6 continental ancestries. This resulted in identification of 6636 individuals of African ancestry and 8876 individuals of Central-South Asian ancestry in the UKBB project. Similar to the above-described case-control definitions, cases were defined using ICD-9/10 codes for atopic dermatitis, excluding other forms of dermatitis, from hospital in-patient data and primary care. Controls excluded individuals with any dermatitis ICD-9/10 codes or AD comorbidities (asthma and allergic rhinitis). The UKBB African ancestry association analysis included 146 cases and 4,799 controls and the Central South Asian ancestry association analysis included 376 cases and 5,594 controls (**Supplementary Table 1**).

GWAS for the UKBB European, African, and Central South Asian cohorts was performed using mixed logistic regression model including sex, age, the first 10 genetic principal components and a genetic relatedness matrix. To fit the model we used SAIGEgds, an R package that implements the Scalable and Accurate Implementation of GEneralized mixed model (SAIGE) method on a Genomic Data Structure (GDS) file format to optimize computational efficiency. SNPs with minor allele count (MAC) > 10 and imputation quality score > 0.7 were kept in the association analysis.

#### Children’s Hospital of Philadelphia (CHOP)

The cohort from the Center for Applied Genomics is composed of approximately 85,000 juvenile subjects below 18 years of age and of a diverse set of ancestries. This study was approved by the Institutional Review Board of Children’s Hospital of Philadelphia (IRB# 4886). Informed consent was obtained from all subjects or, if subjects were less than 18 years of age, from a parent and/or legal guardian, with assent from the child if 7 years or older. Subjects were recruited at Children’s Hospital of Philadelphia starting in 2006 and continuing to the present; genotyping of subjects has occurred across the same timeline. Cases were obtained from ICD-9 diagnosis codes for atopic dermatitis (6918, 6918B), excluding individuals with other forms of dermatitis. Controls excluded individuals with any dermatitis ICD-9 codes (690-698).

Subjects in the study were genotyped on multiple versions of Illumina genotyping arrays. These arrays fell into four families (HumanHap 550/610, Infinium Omni 2.5, Infinium OmniExpress, and Infinium Global Screening Array). Chips within families were merged, filtered for genotype missingness (0.05), individual missingness (0.02) and minor allele frequency (0.05). Genotypes were imputed against the TOPMed reference panel using the TOPMed imputation server^48–50^. Imputed genotypes were filtered for imputation quality using the R^2^ metric (>= 0.6). Post filtering imputed datasets were merged on common SNPs. The final dataset contained 41,180,882 variants.

Ancestry was assigned using PCA. Merged genotypes were filtered for minor allele frequency (0.01) and then pruned for linkage disequilibrium using plink (indep-pairwise 500 50 0.05)^51^. The PCA dataset contained 157,203 SNPs. PCA was performed using flash-pca (v2.0)^52^. The first three principal components were visualized using the plot3D function from the rgl library in R v4.2.3. Subjects were grouped based on observed centers of density into European, African, East Asian, South Asian, and Hispanic/American ancestries. Subjects that fell outside of those five groupings were aggregated into a sixth group, designated “Unassigned”. Genotype files for each ancestry were separated, and another ancestry-specific PCA was performed as above to identify any further outliers.

The association analysis included individuals of African ancestry (6917 cases and 11,080 controls), European ancestry (1590 cases and 21,499 controls), East Asian ancestry (219 cases, 692 controls), South Asian ancestry (123 cases, 1045 controls), American ancestry (237 cases, 1884 controls) and unassigned mixed ancestry (869 cases, 4927 controls). GWAS was performed Scalable and Accurate Implementation of GEneralized mixed model (SAIGE v1.1.4) including sex, age, genotyping batch and the first 10 genetic principal components. SNPs with minor allele count (MAC) >= 5 and imputation quality score >= 0.6 were kept in the association analysis.

#### EAGLE Consortium

The EArly Genetics and Lifecourse Epidemiology (EAGLE) Consortium AD meta-analysis published by Paternoster et al in 2015 performed fixed-effect GWAS meta-analysis on ∼21,000 cases and 95,000 controls. Cases were defined as described in the paper^12^. Summary statistics were generated for the discovery cohort using GWAMA^53^ and were downloaded from GWAS Catalog (excludes 23&Me and non-European cohorts), totaling 10,788 cases and 30,047 controls of European ancestry.

#### Biobank Japan

Biobank Japan collaboratively collects DNA and serum samples from 12 medical institutions in Japan and recruited approximately 200,000 patients with a diagnosis of at least one of 47 diseases. Mean age at recruitment is 63 years. Sakaue et al. performed GWAS analysis on 220 phenotypes, including atopic dermatitis. Summary statistics for atopic dermatitis GWAS were downloaded from the Biobank Japan portal (https://pheweb.jp/downloads). The GWAS summary statistics include 4,296 AD cases and 163,807 controls run using SAIGE (v.0.37) and included age, age^2^, sex, age × sex, age^2^ × sex and the top 20 principal components as covariates. Cases were defined by ICD-10 code L20 and controls were individuals without a L20 ICD-10 code in their medical history.

### GWAS Meta-analysis

Multi-ancestry GWAS statistics were obtained by fixed-effect inverse variance weighted meta-analysis of the summary statistics (beta values) from FinnGen, UKBB_EUR, UKBB_AFR, UKBB_CSA, EAGLE, BBJ and the 6 CHOP cohorts, using GWAMA^53^ (**Supplementary Table 1**). Ancestry-stratified GWAS statistics were obtained by performing an analogous meta-analysis strategy considering cohorts stratified by continental populations: EUR, ASN, AFR (**Supplementary Table 2**). In total, GWAS signal from 4 ancestry endpoints – multi-ancestry, EUR, ASN and AFR populations – were generated. Subsequently, we identified autosomal loci with GWAS signal, i.e. genomic windows containing independent GWAS signals, across ancestry endpoints. For that, we first constructed a reference dataset of best-guess genotypes from UK Biobank (UKBB)^54^ by considering imputed dosages of variants with info score > 0.3 and MAF > 0.1%, selecting genotypic data corresponding to 15,000 randomly selected or to 2,000 ancestry-matched unrelated UKB samples, to generate multi-ancestry or ancestry-stratified genotype panels, respectively. We then filtered variants with missingness > 5% and Hardy–Weinberg equilibrium test P<1 × 10−7. For each of the 4 GWASs, we used the PLINK ref ‘clumping’ algorithm to select top-associated variants (P < 5 × 10−8) and corresponding LD-linked variants at r2 > 0.05 with the top associated variant within ±1 Mb, utilizing the GWAS-matching ancestry-stratified or multi-ancestry UKB genotype data. We determined the genomic span of each LD-based clump and added 1 kb up- and downstream as buffer to the region. If any of these windows overlapped, we merged them together into a single (larger) locus.

To determine a set of non-redundant GWAS loci across ancestry endpoints, we selected all multi-ancestry derived clumps, and complemented this set with non-overlapping clumps identified in a single ancestry. The resulting GWAS hit loci set is composed of genomic regions with suggestive GWAS signal in at least one ancestry endpoint. For each GWAS locus, considering the ancestry endpoint were the GWAS hit was identified, the smallest p-value per locus was defined as the proxy GWAS lead variant for that locus. We filtered out GWAS hit loci overlapping the Major Histocompatibility region (MHC); the final set is composed of 101 GWAS loci.

Comparison of the 101 loci with published literature was performed for atopic eczema/eczema (EFO_0000274, HP_0000964, **Supplementary Table 10 and 11**), allergic disease (MONDO_0005271 **Supplementary Table 12**) and asthma (MONDO_0004979, **Supplementary Table 13**) studies reported in the GWAS catalog with lead SNPs P < 5e-6 (https://www.ebi.ac.uk/gwas; 2023-02-17). Additionally, we extracted reported lead SNPs from Pasanen^33^ (**Supplementary Table 14**) and Budu-Aggrey^14^ (**Supplementary Table 15**) AD GWAS publications. Overlap analysis was performed in R with data.table foverlaps() function. For the 101 genomic loci, start and stop positions windows were set to ±0.5 Mb from the lead variant and intersected with lead SNP positions from GWAS catalog, Pasanen^33^ and Budu-Aggrey^14^ studies. Novel AD loci are defined as loci not overlapping previously published loci from GWAS catalog atopic eczema/eczema, Pasanen^13^ or Budu-Aggrey^14^ studies.

### LDSC heritability

LDSC regression (v1.0.1) was applied to estimate the SNP-based heritability (h^2^_SNP_) of AD from the European ancestry GWAS meta-analysis. h^2^_SNP_ was estimated on liability scale using population prevalence (--pop-prev) 0.15 and sample prevalence (--samp-prev) 0.095 in the meta-analyzed sample (4-cohort EUR meta-analysis 42,963 cases/451,435 controls).

### Fine-mapping

Statistical fine-mapping was performed using the SuSiE with GWAS summary statistics from the European, Asian and African meta-analyses and LD reference panels calculated from UKBB for EUR, AFR and EAS individuals as classified by the Pan-UKBB project (pan.ukbb.broadinstitute.org). We defined fine-mapping regions based on a 1 Mb window around each lead variant and excluded the major histocompatibility complex (MHC) region from analysis due LD structure in the region. The model allowed up to 10 causal variants per region and 95% credible sets (CS) were calculated with posterior inclusion probabilities (PIP) of each variant reported. In loci with multiple causal variants identified, there will be multiple 95% CS. All SNPs in the 95% credible sets were annotated with VEP (http://grch37.ensembl.org/Homo_sapiens/Tools/VEP) using default criteria to select one block of annotation per variant.

### LDSC – cell-type specificity

LDSC-SEG (v1.0.1) was used to identify genomic annotations enriched for AD trait heritability. Bulk ATAC-seq data (GSE118189) for cells isolated from peripheral blood from healthy donors was used to test for enrichment in open chromatin regions. These isolated cells were cultured *in vitro* with and without stimulation as described by Calderon et al^22^. Together there are 222 samples for 32 types of immune cells, 20 of which have data for both unstimulated and stimulated status. Each of cell-type/ATAC-seq bed files was added to the baseline model independently when building the regression model and testing for enrichment.

### Generation of cell-type-specific and disease program gene scores

We scored genes for skin cell-type-specific programs by testing for differential expression by cell-type in healthy and lesional AD cells independently, utilizing a sc-RNAseq dataset derived from skin in the Human Cell Atlas^25^. Briefly, sc-RNAseq data of skin biopsies from 5 healthy controls (HC) and 4 atopic dermatitis (AD) patients were analyzed. The skin biopsies were separated into epidermis and dermis before dissociated and enriched for various cell fractions (keratinocytes, fibroblasts, and endothelial cells) and immune cells (myeloid and lymphoid cells) to up sample rare cell-types. In total, the HC group skin samples sc-RNAseq dataset includes 195,739 cells and AD lesion group sc-RNAseq dataset includes 63,512 cells). Cells were clustered using UMAP dimensionality reduction and Leiden graph-based method then annotated by comparing differentially expressed genes between clusters to published bulk transcription profiles or protein expression of defined cell types. Four major groups of cell types (lymphoid cells, myeloid cells, keratinocytes, and other non-immune cells) were identified and further clustered in subsequent rounds of feature selection, embedding, visualization and clustering. To generate cell-type-specific programs, we first identified genes specifically expressed in one cell type compared to other cell types in HC or AD sc-RNAseq datasets. To generate disease progression programs, we identified differential expression between cells of the same type in AD vs. HC datasets. The p-values from gene programs were transformed to values between 0 and 1 using the min-max normalization resulting in a relative weighting of genes in each program.

### Gene-program enrichment analysis

We adapted MAGMA v1.09b to evaluate the association of disease-association statistics with the cell-type-specific gene programs and disease progression gene programs. In step 1, we performed a gene-level analysis to quantify the degree of association each gene with the AD risk GWAS signal. For each gene g the gene p-value p_g_ from step1 was converted to a Z-value z_g_ with the probit function. In step 2, we performed a competitive gene-program analysis where we modeled the generated cell-type-specific and disease program gene scores and tested whether for a particular gene program, the top-scored genes were more associated to AD risk GWAS signal than low-scored genes. Clustering of gene program scores was performed using the R program pheatmap with Pearson correlation (complete) as the clustering distance method for both rows and columns.

### Determination of QTL-AD-GWAS colocalized loci

To investigate possible associations between cis-genetically regulated molecular phenotypes (QTLs) and Atopic Dermatitis (AD), we compiled an exhaustive QTL map collection and employed a colocalization approach.

### Compilation of QTL full-summary statistics maps

To maximize the expectation of identifying Atopic Dermatitis putatively causal molecular links, we compiled an exhaustive collection of cis quantitative trait loci (QTL) mappings (maps) derived from several molecular phenotypes (MPs): gene (eQTLs), splicing phenotypes (sQTLs), DNA methylation (mQTLs) and protein abundance (pQTLs).

The QTLs originate from widely different contexts, i.e., tissue and cell types, stimuli, and developmental states; we considered a total of 297 cis QTL maps with full statistics available and genome-wide molecular phenotype tests. The majority (94%) of QTL maps are derived from gene expression or splice phenotypes (e/sQTLs), 4% are derived from DNA methylation (mQTLs) and 2% from protein (pQTLs) abundances. Details of QTL maps are provided in **Supplementary Table 6**. A total of 157 eQTL maps were obtained from bulk-tissue or isolated cells, 127 of which from 31 different studies included in the eQTL Catalogue (https://www.ebi.ac.uk/eqtl/, version 5, April 2022. Thirty additional bulk-tissue and isolated-cell eQTL maps were obtained from additional sources: two meta-analyzed eQTL maps derived from blood (eQTLGen, https://eqtlgen.org/cis-eqtls.html) and induced pluripotent stem cells (iPSC) (i2QTL, https://doi.org/10.5281/zenodo.4005576), and 28 maps derived from isolated immune cells (ImmuNexUT, https://humandbs.biosciencedbc.jp/en/hum0214-v8#E-GEAD-420). In addition, 14 immune cell eQTL maps derived from single cell RNA-Seq (sc-eQTLs) were obtained (OneK1K, https://onek1k.org/). Considering splicing phenotypes, we included 109 sQTL maps derived from transcript abundances included in the v5 eQTL Catalogue. Considering DNA methylation, a total of 11 mQTL maps were obtained. We included 9 maps from eGTEx sources: breast mammary tissue, colon transverse, kidney cortex, lung, muscle skeletal, ovary, prostate, testis, and whole blood (eGTEx, https://gtexportal.org/home/downloads/egtex), one additional muscle skeletal (FUSION, https://www.ebi.ac.uk/birney-srv/FUSION/) and one brain (ROSMAP, http://mostafavilab.stat.ubc.ca/xQTLServe/) cis mQTL maps. Considering protein abundance, we included six pQTL maps from plasma (SomaScan deCODE 2021, https://download.decode.is/form/folder/proteomics, SomaScan Sun et al. 2018, https://www.ebi.ac.uk/gwas/downloads/summary-statistics; SomaScan and Olink FinnGen https://www.finngen.fi/en/access_results; ARIC EUR and AFR SomaScan http://nilanjanchatterjeelab.org/pwas).

### Colocalization of AD GWAS loci with QTLs

For each of the 101 AD associated loci, we identified overlapping (> 1 bp) molecular phenotype (MP) *cis*-region loci from each QTL map, considering GWAS and molecular phenotype *cis*-QTL analyzed variants. For each overlapping MP-GWAS region pair, we applied *coloc* v5.52 to QTL along with GWAS summary statistics, only if the locus contained >=1 variant with nominal QTL P < 1e-05 and GWAS P < 5e-08. Prior probabilities of a variant yielding a) a QTL association (p1), b) a GWAS association (p2) and c) a QTL and a GWAS association (p12) were set to p1 = 1e-04, *p*2 = 1e-04, *p*12 = 1e-06. Only the regions with at least 50 variants in common between the GWAS and MP loci were tested for colocalization. Both for QTLs and GWAS statistics, colocalization was performed on effect size (effect size) and associated standard error (effect size s.e.) values, except for Immunext eQTLs; colocalization was performed on p-values and minor allele frequency (MAF) values. We defined suggestive support for QTL-AD-GWAS colocalization at posterior probability PP4 > 0.75. For mQTLs, CpG probe identifiers were mapped to genes according to regulatory region annotations from EPIC.hg38.manifest.tsv.gz and HM450.hg38.manifest.tsv.gz from https://zwdzwd.github.io/InfiniumAnnotation.

### Variant-to-gene mapping and prioritization of high-confidence genes

Given GWAS hit loci, candidate genes per locus can be prioritized by combining evidence across molecular resources^55^. Here we performed variant-to gene mapping and aggregated functional annotation from various sources to generate scores that aim to represent the likelihood of a gene to be causally involved in AD. Genes with the highest aggregated score are named ‘prioritized genes’. Variants were mapped to genes using the combination of multiple methods: nearest gene, Open Targets Variant-to-Gene (V2G), DEPICT, fine-mapping and colocalization. Nearest gene for each genome-wide significant loci was annotated by GREAT (version 4.0.4)^56,57^ using the lead SNP, association rule: Single nearest gene: 1,000,000 bp max extension. Open Targets Variant-to-Gene (V2G)^58^ was used to assign lead variants to genes by selecting the gene with highest overall V2G score. DEPICT (Data-driven Expression-Prioritized Integration for Complex Traits))^59^ was run on all variants with P < 5e-08 to prioritize most likely causal genes at associated loci based on functional annotation. We identified 102 genes with significant (FDR < 0.05) DEPICT score, across 59 loci. VEP annotation of fine-mapped credible sets identified 9 genes with moderate or high impact coding variants (**Supplementary Table 4**). And we utilized VEP to annotate 60 genes containing coding variants in LD (r2 >0.6) with corresponding GWAS locus lead (**Supplementary table 5**). Considering colocalization results per GWAS locus, we annotated all genes with at least one significant colocalization (PP4 > 0.75, as described in above colocalization methods) derived from sQTL, eQTL and pQTL maps (**Supplementary Table 7**). Considering deCODE and FinnGen *trans* pQTL summary statistics (source) corresponding to GWAS loci lead SNPs, we annotated genes with *trans* pQTL signal. That is, if lead SNP of GWAS locus 1 had a nominally significant (P <1e-05) *trans* pQTL signal associated to a protein encoded in GWAS locus 2, the GWAS locus 2 gene encoding such protein would be annotated with *trans* pQTL signal.

All genes implicated by any of the above strategies were then annotated with OMIM entries associated to reported phenotypes involving the skin or immune system involvement (**Supplementary Table 8**). With this approach, we annotated 57 genes with a potential role in skin or immune processes. Additionally, genes reported in literature with coding variants in AD patients were compiled and annotated against the gene list (see reference publications in **Supplementary Table 16**) And finally, genes with significant differential expression (FDR < 0.01, log2FC > [1]) from Tsoi et al 2019^31^ comparisons of AD lesional skin to control (healthy) skin bulk RNAseq were annotated (n=3264 DEGs, see **Supplementary Table 17**). The amount of evidence across all sources was added in an unweighted fashion to generate aggregated scores for 498 unique genes across 101 GWAS loci (**Supplementary Table 8**).

### Selection and prioritization of AD keratinocyte-linked gene candidates

We selected genes with =>1 s/eQTL colocalization signal (PP4 > 0.75), where the AD risk allele increases the expression of the gene or transcript in at least one s/eQTL endpoint. We integrated skin cell enrichment metrics (Supplementary Table 2 of Dusart et al.^60^) and narrowed down this set by selecting genes with evidence of correlation with keratinocyte-representative transcripts, e.g. with mean correlation with keratinocyte reference transcripts > 0.30, and that being higher than the mean correlation with any non-keratinocyte reference transcript set. The selected set is composed of 22 genes: *AQP3, NAB1, CEBPA, IL2RB, RORA, GRID2IP, RGS14, RTF1, LIME1, ZFYVE21, SLC2A4RG, SCAMP3, IL22RA2, WNK1, CLIP1, KIAA2013, LMAN2, MAP3K14, NDUFA4L2, ANK3, CHRAC1, PCDH1.* We refer to this set as “AD keratinocyte-linked gene candidates”; genes that are candidates to play a causal role in Atopic Dermatitis through a pathogenic effect in keratinocytes.

### Characterization of keratinocyte subtype specificity signal using a cross-body sc-RNA-Seq skin dataset

To assess the robustness of the keratinocyte-specific gene expression of the keratinocyte-linked colocalized genes, we generated a sc-RNAseq dataset composed of AD unaffected epidermal samples from 96 skin biopsies from seven body sites (face, scalp, axilla, palmoplantar, arm, leg, and back). The skin biopsies were separated into epidermis and dermis before dissociated and enriched for various cell fractions (keratinocytes, fibroblasts, and endothelial cells) and immune cells (myeloid and lymphoid cells) to up sample rare cell types. In total, across body sites, 274,834 cells were profiled, including 96,194 keratinocytes. *Seurat v3.0.* was utilized to normalize, scale, and reduce the dimensionality of the data. We filtered out low quality cells containing less than 200 genes per cell as well as greater than 5,000 genes per cell. Cells containing more mitochondrial genes than the permitted quantile of 0.05 were removed. We removed ambient RNA using R package *SoupX* v1.6.2. Doublets were removed using *scDblFinder* v1.12.0. Principal components (PC) were obtained from the topmost 2,000 variable genes, and the Uniform Manifold Approximation and Projection (UMAP) dimensional reduction technique was applied to the 30 topmost variable PC-reduced dataset. Batch effect correction was performed utilizing *harmony* v1.0, using donor as batch. After batch correction, cells were clustered using shared nearest neighbor modularity optimization-based clustering. Cluster marker genes were identified with *FindAllMarkers*; cluster corresponding cell type was identified by comparing marker genes to curated cell-type signature genes (**Extended Figure 6**). Differential expression by keratinocyte subtype was performed with Seurat (v4.3.0) *FindMarkers* function by comparing keratinocyte subtype to non-keratinocyte clusters. The log fold-change of the average expression between a keratinocyte subtype cluster compared to the rest of clusters is utilized as keratinocyte-subtype gene expression statistic. Biopsy sample details, raw data, processed Seurat object, and curated cell-type signature genes are available in SRA (id PRJNA1054546) and FigShare (https://plus.figshare.com/account/home#/projects/198796).

### Characterization of keratinocyte-differentiation gene expression signal using 3-D human epidermal tissue cultures

We investigated the gene expression patterns of the keratinocyte-linked colocalized genes in keratinocyte differentiation occurring in the formation of 3-D human epidermal raft cultures. Normal human epidermal keratinocytes were isolated from epidermis (n=3) and grown using J2-3T3 mouse fibroblasts as a feeder layer originally described by Rheinwald and Green^61^. 3-D human epidermal raft cultures seeded in collagen hydrogels were prepared using three distinct donor pools as described previously^62^ and grown at an air-liquid interface for 12 days in E-Medium (DMEM/DMEM-F12 (1:1), 5% Fetal Bovine Serum, adenine (180µM), Bovine pancreatic insulin (5µg/ml), Human apo- transferrin (5µg/ml), triiodothyronine (5µg/ml), L-Glutamine (4mM), Cholera toxin (10ng/ml), Gentamicin (10µg/ml), Amphotericin B (0.25µg/ml)). At day 9 at an air-liquid-interface to allow for epidermal maturation, the epidermal rafts (RHE) were treated with 0.1% BSA/phosphate-buffered saline (Sigma Aldrich, St Louis, MO) for 72 Hrs. Epidermal tissues were separated at the stages from Sub-confluent stage to 3-D raft on day 12 (Sub-confluent, Day 0-Confluent, Day 3-Confluent, Day 3-Raft, Day 6-Raft, Day 9-Raft, Day 12-Raft) from the collagen scaffold and lysed in QIAzol for RNA isolation. RNA samples were sent to the University of Michigan Advanced Genomics Core for RNA sequencing. Libraries for RNA-Seq were generated from polyadenylated RNA and sequenced at six libraries per lane on the Illumina Genome Analyzer IIx. We used Tophat2^63^ to align RNA-seq reads to the human genome, using annotations of GENCODE as gene model^64^. HTSeq was used to quantify gene expression levels^65^. Normalization was performed by DESeq2^66^. The processed RNA-Seq data can be found in FigShare (https://plus.figshare.com/account/home#/projects/198796). Differential expression by timepoint was performed with *limma*^67^ on FPKM values on non-lowly expressed genes (zFPKM > -3, in at least 2 timepoints). Timepoint was modeled as a quantitative variable (Sub-confluent=1, Day 0-Confluent=2, Day 3-Confluent=3, Day 3-Raft=4, Day 6-Raft=5, Day 9-Raft=6, Day 12-Raft=7), and the function *duplicateCorrelation* was utilized to model technical replicate effects. The estimated log-fold change attributable to timepoint is utilized as keratinocyte-differentiation gene expression statistic.

### Characterization of candidate gene silencing in IL-13 and IL-22 pathways

To characterize the effects of silencing AD keratinocyte-linked genes on IL-13 and IL-22 pathways, which are implicated in AD pathogenesis^40,68^, we knocked-out (KO) candidate genes with silencing RNAs (siRNA) in N/TERTs^69^ immortalized keratinocytes cells, and evaluated the expression of interleukin pathway proxy genes S100A9, S100A8, S100A7 (IL-22) and CCL26, CISH, HSD3B1 (IL-13). Keratinocytes were plated in 96 well plate (20,000 cells/well) and incubated at 37°C with 5% CO2 overnight. 100μM Accell siRNAs for AD keratinocyte-linked gene candidates (**Supplementary Table 9**) were prepared in 1x siRNA buffer (Dharmacon# B-002000-UB-100). 1μl of 100μM siRNA was diluted with 100μl accel delivery medium (Dharmacon # B-005000) for each well of 96 well plate. The growth medium was removed from the cells, 100μl of the delivery mix with siRNAs was added to each well, and the plate was incubated at 37°C with 5% CO_2_. Accell Non-targeting Control siRNA (Dharmacon # D-001910-01-05) was used as a negative control. After 24 hours, cells were either stimulated with 10 ng/ml of IL-13 (R&D Systems # 213-ILB), 20 ng/ml of IL-22 (R&D Systems # 782-IL) or co-stimulated with IL-13 (10 ng/ml) and IL-22 (20 ng/ml). After 24 hours of stimulation, cells were harvested for RNA preparation. RNAs were isolated from cell cultures using Qiagen RNeasy plus kit (Cat # 74136). Reverse transcription was performed using a High-Capacity cDNA Transcription kit (ThermoFisher # 4368813). qPCR was performed on a QuantStudio 5 Real-time PCR system (Applied Biosystems) with TaqMan Universal PCR Master Mix (ThemoFisher # 4304437) using TaqMan primers. *RPLP0* (ThermoFisher # Hs99999902_m1) was used as a loading control. Three technical replicates were considered. Knockdown efficiency was validated by the TaqMan primer of each of the siRNA target genes (**Supplementary Table 18**). KO efficiency values are illustrated in **Extended Fig. 9**. The protocol failed for LIME1 and MAP3K14, which were not considered in further analyses. The differential expression of proxy genes between the presence or absence of siRNA targeting corresponding gene candidate was evaluated in each condition by t-test, considering the three technical replicates, and derived standardized mean difference (SMD) effect size and corresponding sampling variance were generated with f(x) ’escalc’ from ’metafor’ v.4.0. SMD values were meta-analyzed across markers per pathway, considering results from IL-13+IL-22 treatment, with *metafor::rma* function.

### Mapping of keratinocyte eQTLs

We mapped eQTLs in keratinocyte cell lines derived from N=50 subjects, for which RNA-Seq profiles and genotype data were generated^70^. Cell lines were subjected to 8 different conditions implicated in AD pathogenesis^31,71^: no stimulation, TNF (10ng/ml), IL-4 (10ng/ml), IL-13 (10ng/ml), IL-17A (10ng/ml), IL-17A+TNF, IFNa (5ng/ml), IFNg (5ng/ml). Gene expression values were first normalized by DESeq2, and PEER was used to account for latent confounding factors. The genotype data was generated by the Illumina Infinium CoreExome array, and imputation was performed using 1000 Genomes Project (GRCh37/hg19) as reference panel. Cis (+/- 1Mb from gene transcription start site) eQTL were mapped using *FastQTL* v2.0 by fitting a linear regression model (p ∼ g + **C**) where p is the gene expression vector, g is the genotype vector, and **C** is a matrix of 10 PEER factors derived from gene expression; eQTLs signal was assessed by the effect size corresponding to the term g. Full summary statistics are provided in https://plus.figshare.com/account/home#/projects/198796. The eQTL mappings restricted to 22 variant-gene pairs corresponding to AD keratinocyte-linked gene candidates - 20 AD GWAS loci and 22 genes - are provided in **Supplementary Table 9**. These eQTL effects were estimated considering the index variant of the AD GWAS hit corresponding to the keratinocyte-linked colocalized gene.

### Correlation of keratinocyte assays’ readouts with candidate priority status

The set of 22 keratinocyte-linked genes was profiled by four different functional assays in keratinocytes (see corresponding Method sections above). We hypothesized that keratinocyte-linked gene candidates prioritized as more likely to be causal would yield more signal across assays, indicative of their active role in key keratinocyte pathways. To test this hypothesis, we first classified 5/22 candidates (*AQP3, CEBPA, RORA, RGS14, ANK3)* as ‘prioritized’, by having top aggregated scores per locus and accounting for more than half of eQTL colocalization instances per locus (**Supplementary Table 9, Fig. 5**). We then compared aggregated corresponding assay readouts with the remaining 17/22 non-prioritized candidates (**Extended Fig. 7**) and assessed significant differences in statistics distribution by means of Mann-Whitney test. Indeed, the statistics for prioritized genes are significantly larger than for non-prioritized genes in all assays: in IL-13 and IL-22 pathways gene silencing assays (Mann-Whitney P = 0.01 on absolute SMD values), in keratinocyte eQTLs (Mann-Whitney P = 3.58e-15 on absolute eQTL effect size values), in keratinocyte-differentiation gene expression signal (Mann-Whitney P = 1.18e-03 on absolute log-fold change values) and in keratinocyte subtype specificity signal (Mann-Whitney P = 2.82e-06 on absolute log-fold change values).

## Data Availability

We deposited the full GWAS summary statistics (MULTI, EUR, ASN, AFR) to the GWAS Catalog under accession IDs: TO BE DEPOSITED UPON ACCEPTANCE. Additional data files are available in supplementary tables and at FigShare site: https://plus.figshare.com/account/home#/projects/198796 (TO BE MADE PUBLIC UPON ACCEPTANCE OR EDITORS REQUEST)

## CODE AVAILABILITY

The code is available at AbbVie internal github repository TO BE MADE PUBLIC UPON ACCEPTANCE

## ACKNOWLEDGMENTS

We thank Xiuwen Zhang and John Lee for support of UKBB data analysis. We acknowledge Jacob Degner, Emily King, Jeff Waring, and Zoltan Dezso for critical review of the manuscript. This research was carried out using the UK Biobank resource under application number 26041. We acknowledge the Biobank Japan Project for access to summary statistics. We want to acknowledge the participants and investigators of FinnGen study. The FinnGen project is funded by two grants from Business Finland (HUS 4685/31/2016 and UH 4386/31/2016) and the following industry partners: AbbVie Inc., AstraZeneca UK Ltd, Biogen MA Inc., Bristol Myers Squibb (and Celgene Corporation & Celgene International II Sàrl), Genentech Inc., Merck Sharp & Dohme Corp, Pfizer Inc., GlaxoSmithKline Intellectual Property Development Ltd., Sanofi US Services Inc., Maze Therapeutics Inc., Janssen Biotech Inc, Novartis AG, and Boehringer Ingelheim. Following biobanks are acknowledged for delivering biobank samples to FinnGen: Auria Biobank (www.auria.fi/biopankki), THL Biobank (www.thl.fi/biobank), Helsinki Biobank (www.helsinginbiopankki.fi), Biobank Borealis of Northern Finland (https://www.ppshp.fi/Tutkimus-ja-opetus/Biopankki/Pages/Biobank-Borealis-briefly-in-English.aspx), Finnish Clinical Biobank Tampere (www.tays.fi/en-US/Research_and_development/Finnish_Clinical_Biobank_Tampere), Biobank of Eastern Finland (www.ita-suomenbiopankki.fi/en), Central Finland Biobank (www.ksshp.fi/fi-FI/Potilaalle/Biopankki), Finnish Red Cross Blood Service Biobank (www.veripalvelu.fi/verenluovutus/biopankkitoiminta) and Terveystalo Biobank (www.terveystalo.com/fi/Yritystietoa/Terveystalo-Biopankki/Biopankki/). All Finnish Biobanks are members of BBMRI.fi infrastructure (www.bbmri.fi). Finnish Biobank Cooperative -FINBB (https://finbb.fi/) is the coordinator of BBMRI-ERIC operations in Finland. The Finnish biobank data can be accessed through the Fingenious^®^ services (https://site.fingenious.fi/en/) managed by FINBB. JEG, MKS, and LCT are supported by NIH-P30-AR075043.

## CONTRIBUTIONS

B.R.G. and M.O. conceived the study with contributions of M.K.S., M.E.M., H.H., J.E.G. and K.M.S. B.R.G. conceived and performed the meta-GWAS and derived downstream analyses, M.O. conceived and performed QTL colocalization and derived downstream analysis. F.T. conceived and performed the cell-type enrichment analysis, including ideation of novel MAGMA approach. B.R.G. and M.O. conceived and performed candidate gene assessment analysis. M.E.M., supervised by H.H., generated the CHOP GWAS and D. W., J.T.G, A.H.S and F.D.M contributed with QC and interpretative analyses. M.K.S., supervised by J.E.G., conducted the siRNA and 3-D epidermal model experiments. C-L.H. contributed to the profiling of keratinocyte-linked genes to the interpretation of the siRNA experiments. R.B., J.M.K., supervised by L.C.T., generated the keratinocyte eQTLs, and the skin sc-RNAseq experiment and corresponding data. R.U., M.T.P, and Q.L. contributed to quality control, and interpretation of the keratinocyte and skin datasets. B.R.G. and M.O. led the writing and editing of the manuscript and supplement; all authors contributed to the editing of the manuscript and supplement. B.R.G. and M.O. coordinated analyses of all contributing authors. All authors read and approved the final manuscript.

## ETHICS DECLARATIONS

B.R.G, M.O., C.H., F.T., K.M.S are or were employees of AbbVie participated in the design, study conduct, interpretation of data, review, and approval of the publication. JEG has received research support from AbbVie, Janssen, Almirall, Prometheus Biosciences/Merck, BMS/Celgene, Boehringer Ingelheim, Galderma, Eli Lilly, and advisor to Sanofi, Eli Lilly, Galderma, BMS, Boehringer Ingelheim The remaining authors declare no competing interests.

## EXTENDED FIGURES

**Extended Figure 1.**
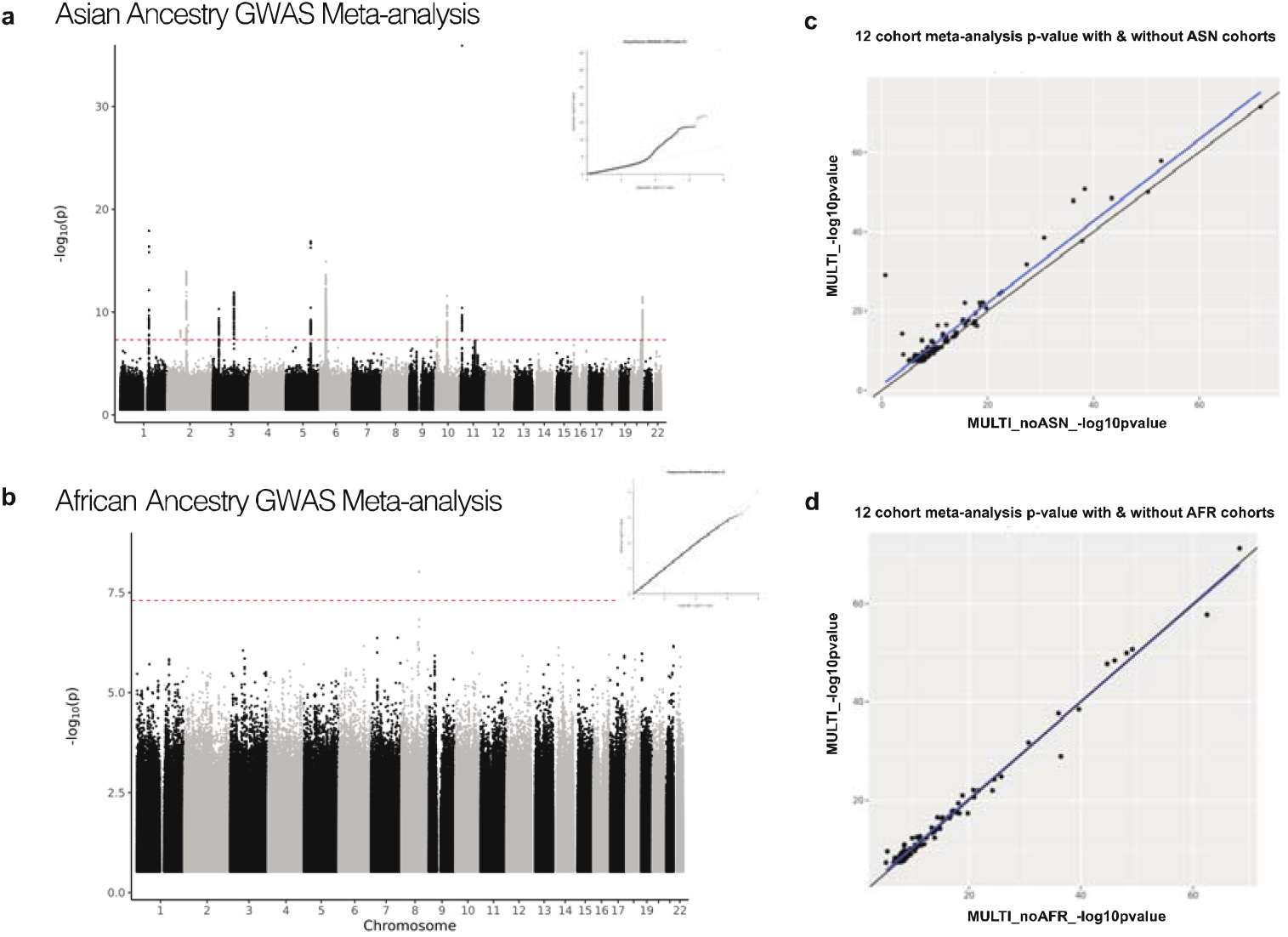
Ancestry-specific GWAS meta-analysis and comparison plots. **a-b** Manhattan and QQ plots derived from ASN and AFR GWAS meta-analysis (Methods). Genome-wide significance threshold (P< 5e-08) is indicated by red dashed line. At MAF > 0.01, twelve significant loci were identified in ASN GWAS (a) and none in AFR GWAS (b). **c-d**, Comparison of the significance of GWAS loci lead variants identified in the MULTI GWAS, with (y axis) and without (x axis) the ASN (c) and AFR (d) cohorts.

**Extended Figure 2.**
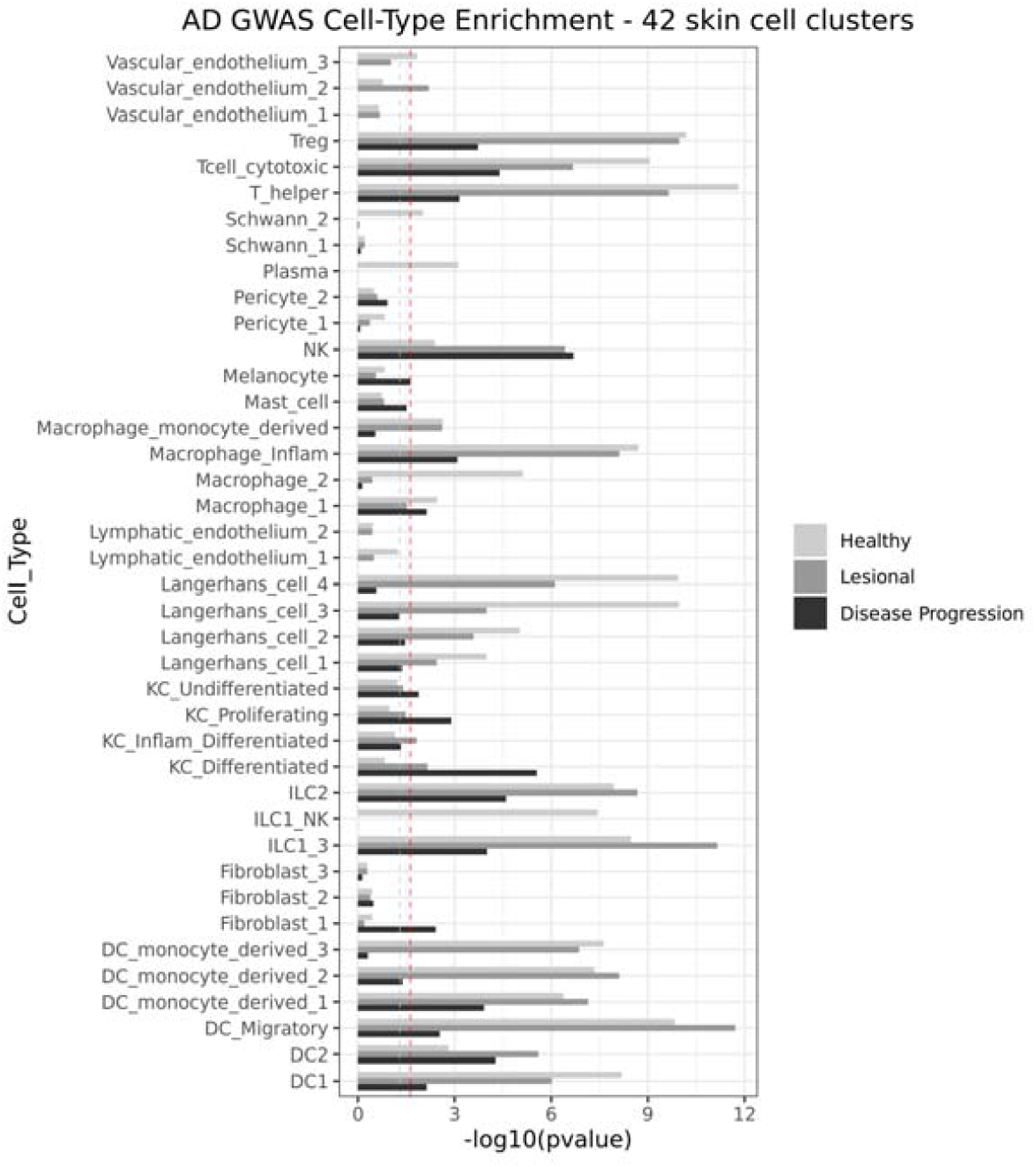
Cell-type enrichment of AD GWAS signal in skin cell types. Cell-type enrichment significance (x axis) for 42 skin cell types (y axis) of AD GWAS signal, derived from HCA human skin sc-RNAseq data. The p-values correspond to the MAGMA enrichment of AD GWAS genes in cell-type-specific gene programs (Methods). Enrichment of cell-type programs of healthy skin and AD lesion skin for AD traits are displayed in light and dark grey, respectively. Enrichment of disease progression programs in AD lesion relative to healthy tissues (LS_vs_H) for AD traits are displayed in black. Dashed grey line is the threshold for p-value < 0.05, and solid red line is the threshold for FDR < 0.05.

**Extended Figure 3.**
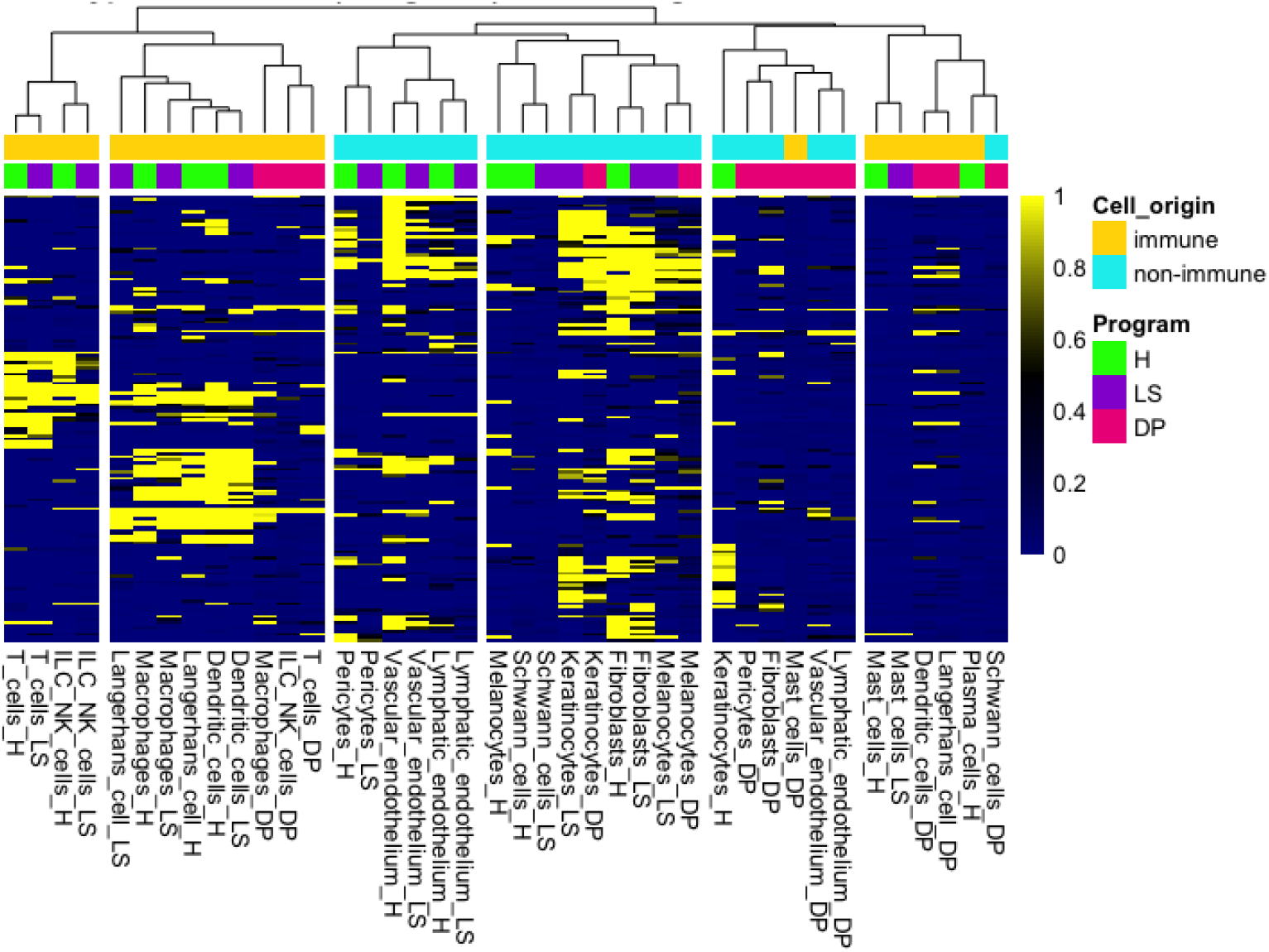
Cell-type enrichment clustering of gene program scores. Gene program scores (0-1) corresponding to significant genes (MAGMA P <2.7e-06) derived from MAGMA analysis (Methods), including 146 genes (y axis) and 14 cell-type programs (x axis). Genes scores were clustered based on Pearson correlation-based distance and complete-linkage. Cell of origin lineage is indicated by yellow (immune cells derived from myeloid and lymphoid lineage) and blue (non-immune cells). Healthy (H), lesional (L), and disease progression (DP) programs are indicated by green, purple, and magenta, respectively.

**Extended Figure 4.**
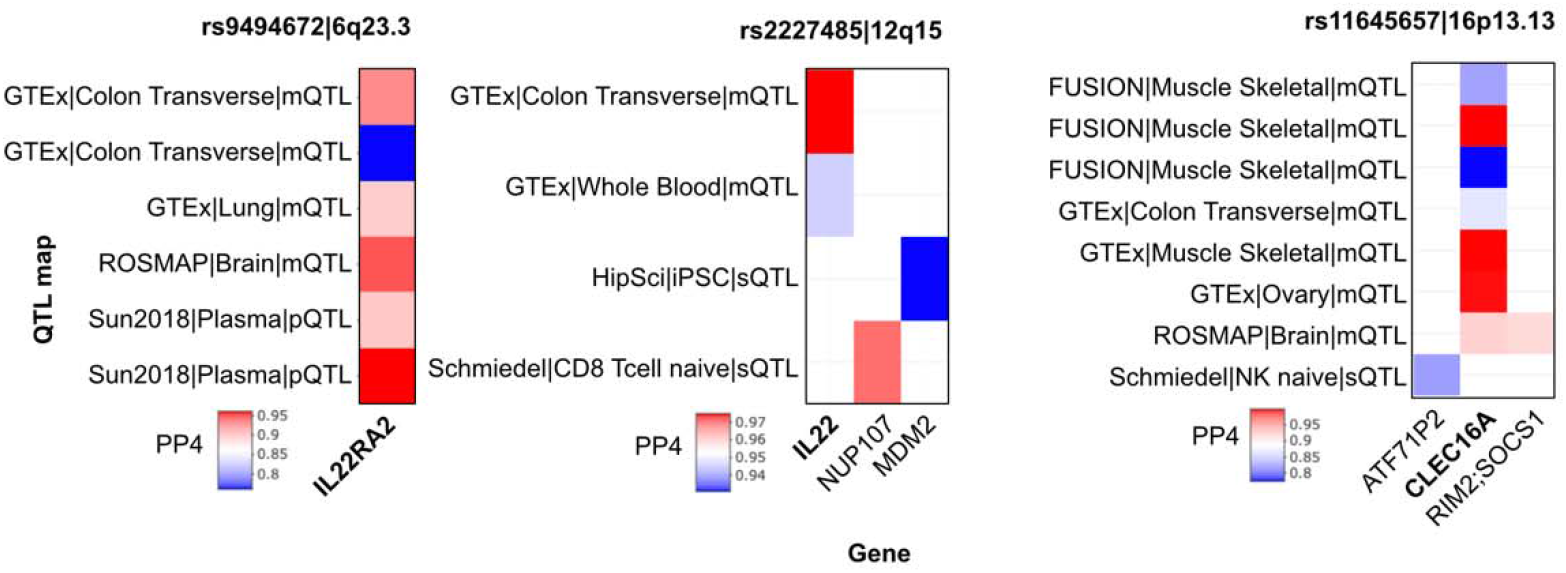
QTL-GWAS colocalizations derived from non-eQTL maps. Colocalization probabilities (PP4) are stratified per AD GWAS locus, QTL map (y axis) and gene (x axis). Genes with the largest number of colocalizations per locus are typed in bold. Each locus is labeled with corresponding lead variant and cytoband.

**Extended Figure 5.**
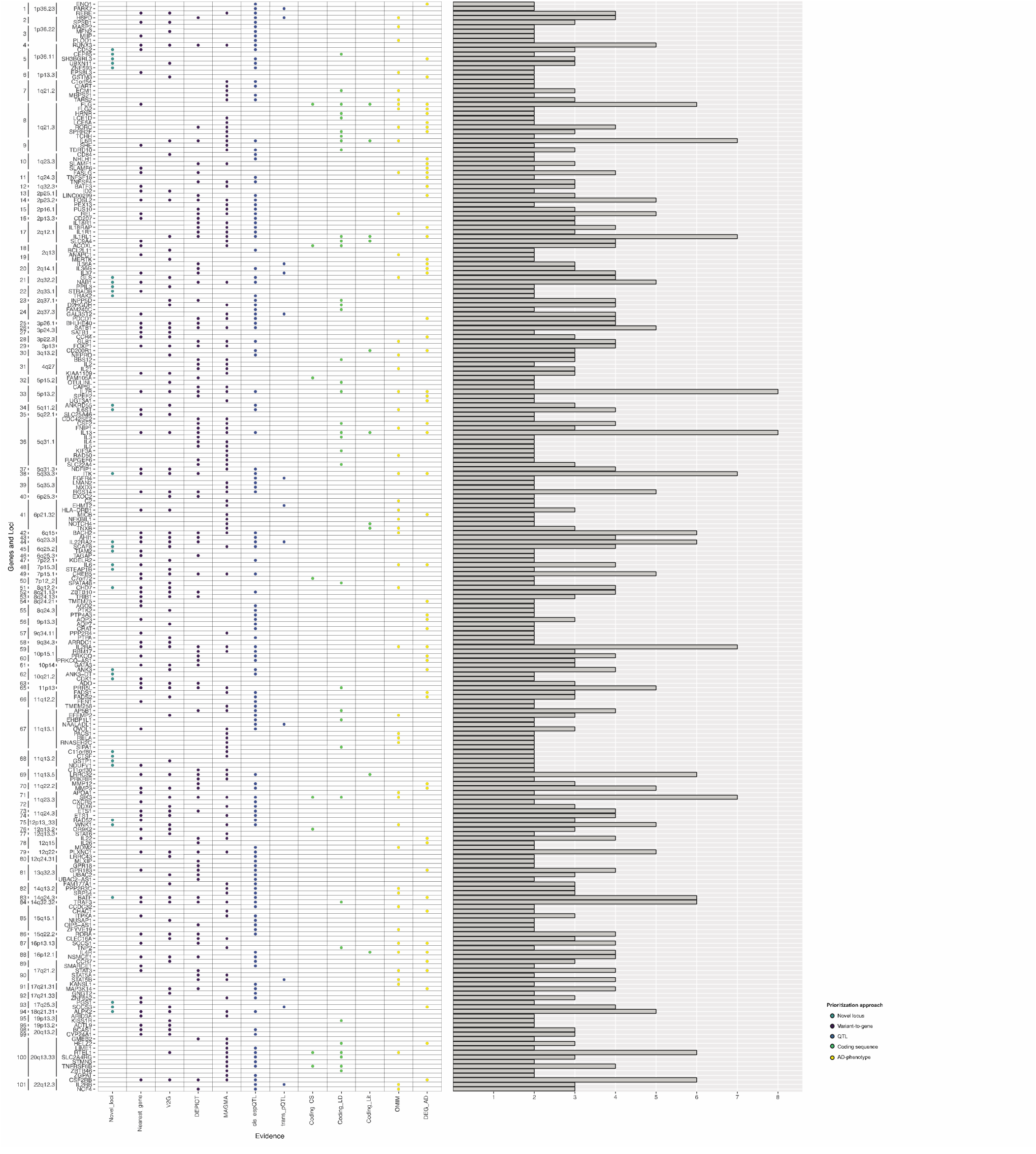
Prioritization scores for genes within AD GWAS loci. Scores derived from annotations (x axis) for genes located within the 101 AD GWAS significant loci (y axis), colored by annotation category (Methods). Barplot illustrates the aggregated sum of scores. Out of 498 scored genes (Supplementary table 8), displayed are 239 genes with ≥2 lines of supporting evidence.

**Extended Figure 6.**
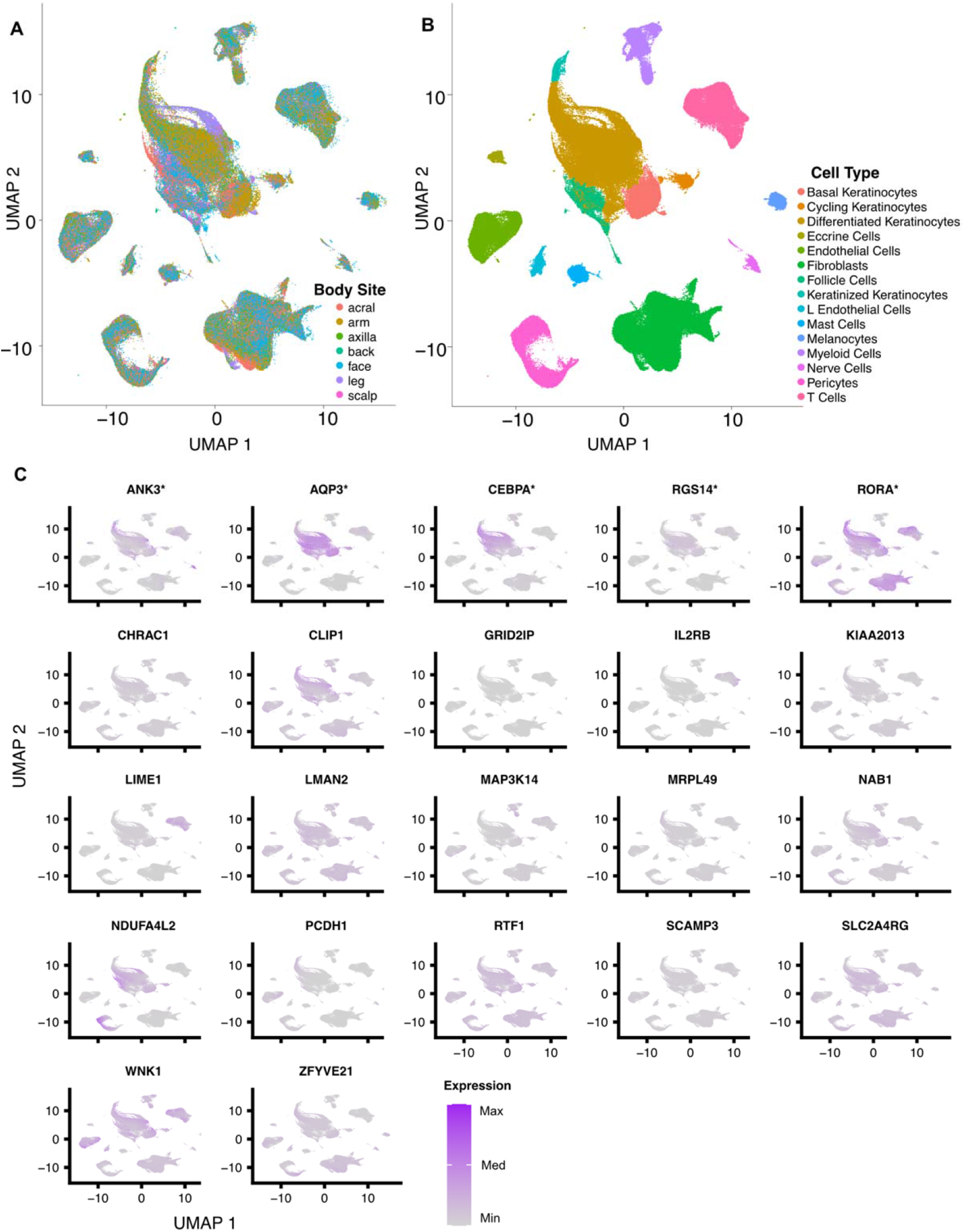
UMAP embedding of sc-RNAseq data for skin samples from seven body sites. In **A**, colors represent sampled body sites. In **B**, colors represent cell types. In **C**, purple color gradient corresponds to the degree of expression of AD keratinocyte-linked gene candidates, scaled by gene. Prioritized candidates (Methods) are indicated with wildcard.

**Extended Figure 7.**
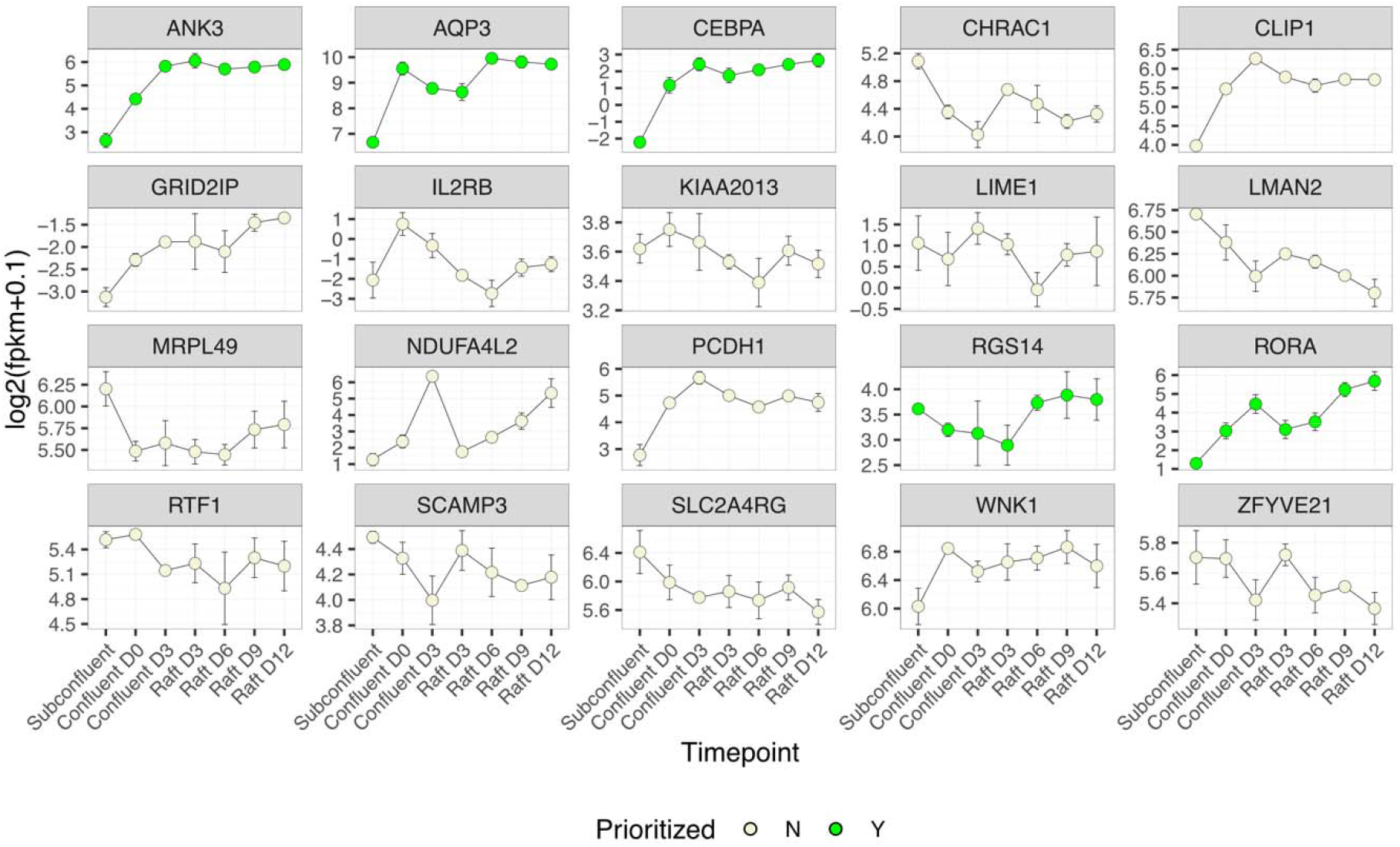
AD keratinocyte-linked gene candidates’ expression by keratinocyte differentiation. Gene expression (y axis) by keratinocyte differentiation timepoint (x axis). Whiskers represent the standard deviation of the mean. Target genes are stratified by prioritization score (Methods); prioritized candidates are colored in green.

**Extended Figure 8.**
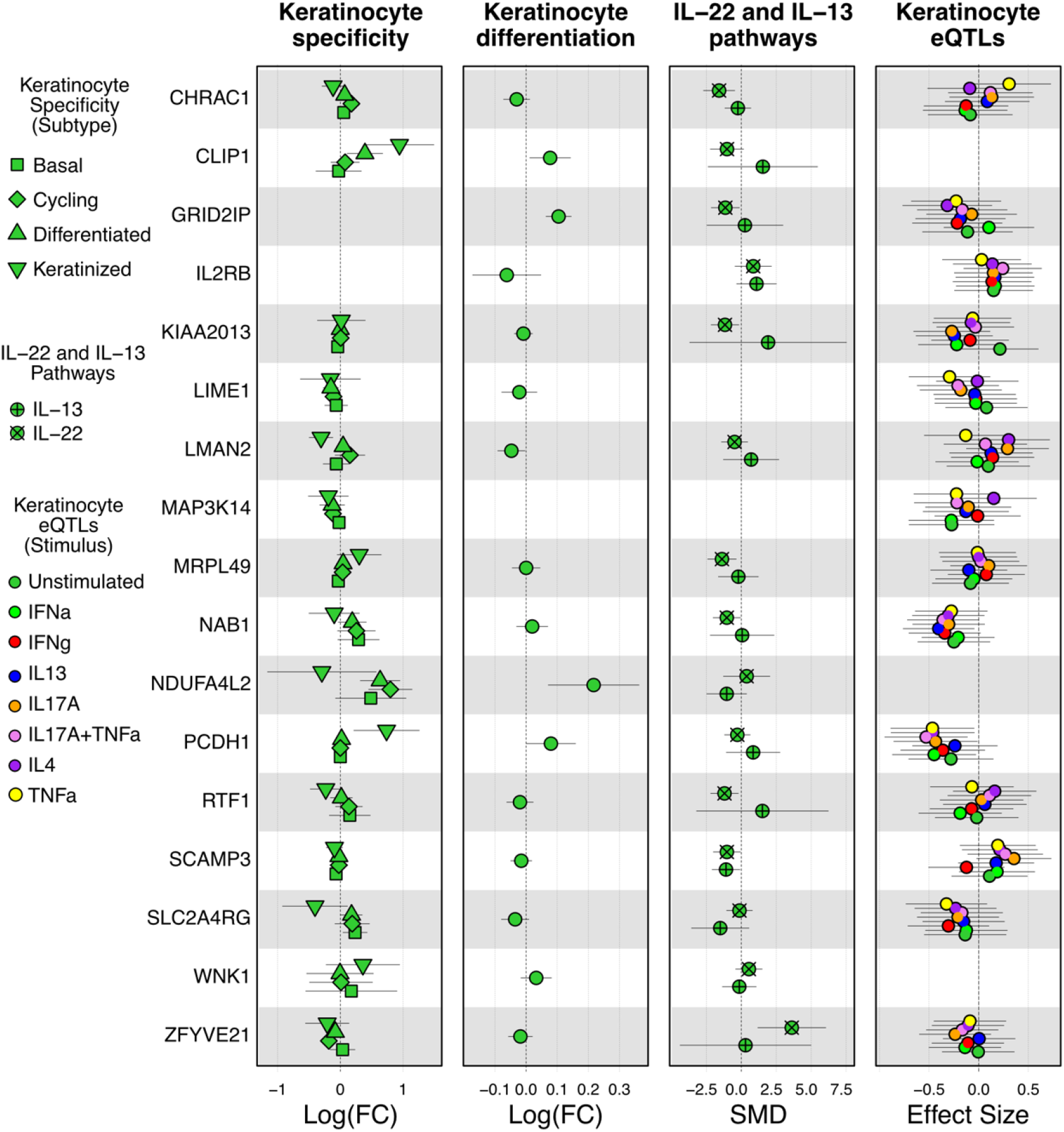
Functional characterization of non-prioritized AD keratinocyte-linked gene candidates. **1st panel**, Keratinocyte subtype differential expression (x axis) by gene (y axis). Differential expression values correspond to expression log fold change (FC) between keratinocyte and non-keratinocyte cells, mean-averaged across body sites, and whiskers represent the standard deviation of the mean. **2nd panel**, Differential expression (x axis) of gene (y axis) as a function of raft differentiation. Differential expression values correspond to log expression fold change as a function of differentiation timepoints (Methods), and whiskers represent the 95% confidence interval of the value. **3rd panel**, Differential expression of pathway (x axis) proxy genes by candidate gene (y axis). Differential expression values correspond to the standardized mean difference (SMD) of the expression of genes that are proxies of IL-22 and IL-13 pathways between the presence or absence of siRNA targeting corresponding gene candidate. Whiskers represent the 95% confidence interval of the value. SMD values were derived from across-proxy-genes per-pathway meta-analysis (Methods). **4th panel**, Keratinocyte eQTL effect size (x axis) by candidate gene (y axis), stratified by keratinocyte stimulus. Effects are mapped to AD risk alleles of the corresponding GWAS locus lead variants. Whiskers represent the 95% confidence interval of the value.

**Extended Figure 9.**
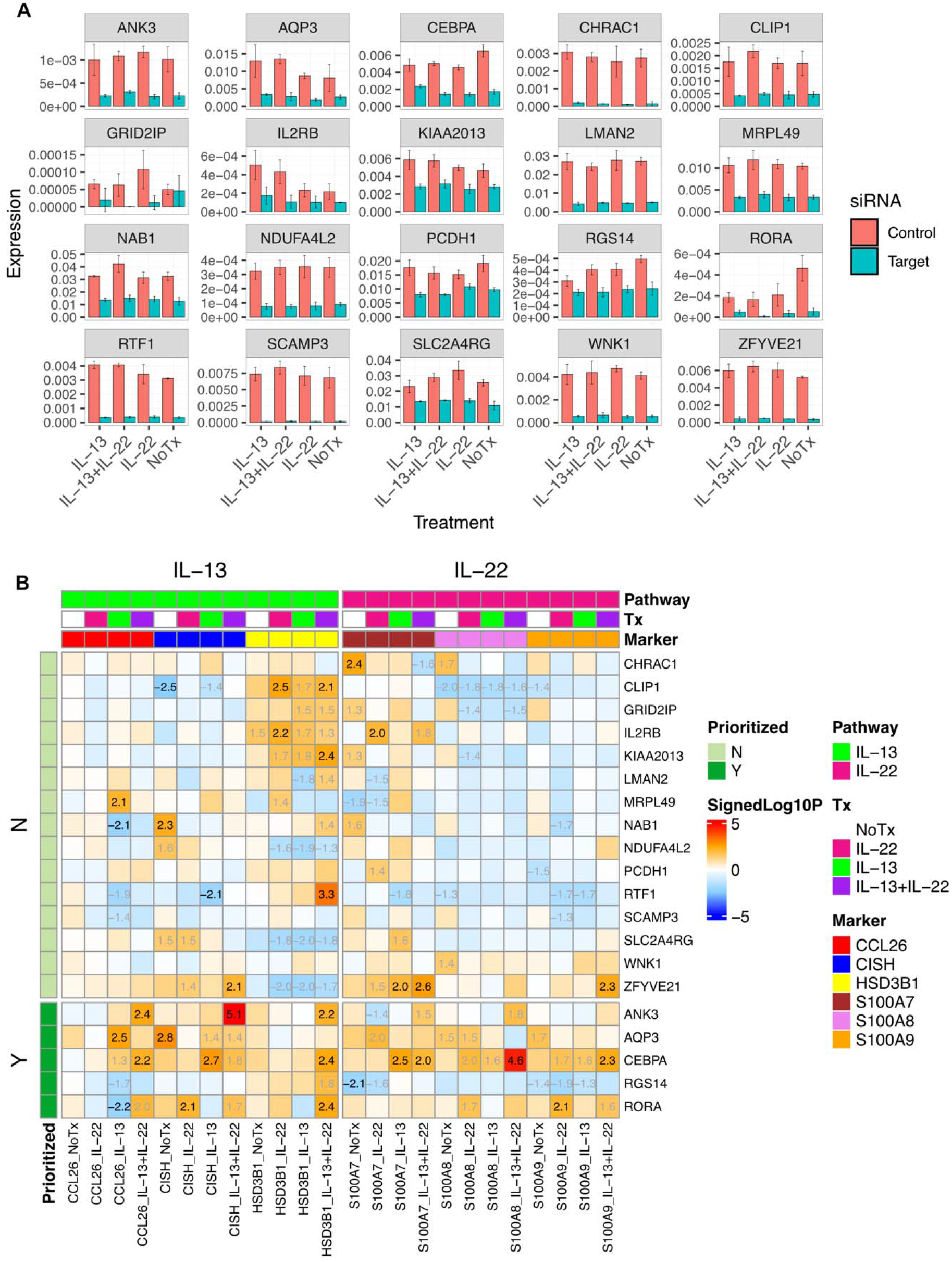
IL-13 and IL-22 impact upon KO of AD keratinocyte-linked gene candidates. **A,** Target gene expression (y axis), stratified by control and target siRNA experiment, for each treatment condition (x axis). **B**, Significance of KO effects of silenced candidate target gene (y axis) on interleukins (IL) IL-22 and IL-13 proxy marker genes (x axis). For each candidate target gene, IL marker gene expression was compared between target KO and NTC (non-target control) experiments, in each treatment (Tx) condition. Differential expression was calculated by means of t-test, considering the mean of 3 technical replicates per experiment. Heatmap values represent the signed -log10 p-values (P) of the corresponding t-test, colored by significance; P<0.01 and 0.01<P<0.05 tests are typed in black and grey, respectively. Values for P>0.05 tests are not shown. Target genes are stratified by prioritization score (Methods).

